# T1w/T2w ratio suggests reduced intracortical myelin content in youth with RASopathies

**DOI:** 10.1101/2025.09.16.25335916

**Authors:** Julia R. Plank, Sara K. Pardej, Mira M. Raman, Jennifer McNab, Tamar Green

## Abstract

**Background:** Myelin may represent a modifiable treatment target in neurodevelopmental disorders, however, a reliable tool for *in vivo* assessment of myelin alterations in clinical settings is needed. Prior work shows commonly acquired T1-weighted (T1w) and T2-weighted (T2w) scans can be transformed into ratio maps and subsequently provide a reasonable estimate of cortical myelin content. We investigated T1w/T2w ratios for measuring cortical myelin in children with neurodevelopmental disorders of the RAS-MAPK signaling pathway.

**Methods:** In this prospective study, 112 children (86 RASopathies, 26 typical developing (TD), aged 6-17 years) completed T1w and T2w MRI scans and NIH Toolbox cognitive assessments. Parent-rated mobility and strength impact scores were also acquired. T1w/T2w ratio maps were calculated at three levels of the cortex in FreeSurfer, and average values were extracted from 68 regions-of-interest (ROIs) at each level across the brain and from eight subcortical ROIs. Group differences were assessed using analyses of covariance, including a false discovery rate adjustment for multiple comparisons. As an exploratory analysis, differences in cognitive scores and parent-rated health were assessed across quartiles of whole-brain T1w/T2w ratios.

**Results:** Widespread decreases in T1w/T2w ratios were found in the RASopathies group, suggesting decreased cortical myelin content. Of the T1w/T2w ratios sampled along the midline, 63 of 68 cortical ROIs were significantly reduced in RASopathies (*p_FDR_*<.050). Of the subcortical ROIs, only the T1w/T2w ratio in the accumbens was significantly reduced in RASopathies compared to TD (Cohen’s *d*=0.520, *p_FDR_*=.024). Exploratory quartile analyses across the whole sample indicated significant effects of T1w/T2w ratio quartile on mobility (*p*=.002) and strength impact ratings (*p*<.001), such that subjects in higher T1w/T2w ratio quartiles had better physical health ratings.

**Conclusions:** These results support the utility of T1w/T2w ratio mapping as a sensitive tool for detecting cortical myelin alterations in genetically defined neurodevelopmental disorders. Exploratory analyses suggest a relationship between cortical myelin content and physical mobility and stamina. Future work should explore the clinical relevance of these findings for cognitive and functional outcomes and assess their potential as biomarkers for targeted therapeutic interventions.

## Introduction

Myelin is an essential component of healthy brain function, supporting efficient communication between brain regions. Changes in myelin content occur throughout the lifespan, typically following an inverted U-shape trajectory with increases in childhood followed by declines in adulthood (1). Abnormalities in myelin structure or development are implicated in neurodevelopmental disorders, and may be linked to cognitive, motor, and behavioral impairments (2). Given the known importance of myelin in brain functioning, myelin is increasingly recognized as a potential treatment target in clinical trials (3). The plasticity of myelin (i.e., its ability to change in response to experience) poses myelin as an ideal candidate for interventions (4). However, a robust, non-invasive tool is needed to establish myelin as a measurable biomarker for translation to human clinical trials.

Magnetic resonance imaging (MRI) is a non-invasive tool used widely for *in vivo* measurement of alterations to brain structure. Recent studies have tested a variety of MRI techniques with the aim of measuring brain microstructure and myelin content. Prior studies suggest calculation of a ratio between T1-weighted (T1w) and T2-weighted (T2w) images may provide a reasonable estimate of intracortical myelin content across the brain (5). By dividing the intensity values from T1w images (which are sensitive to fatty tissues like myelin) by those from T2w images (sensitive to water), the ratio enhances the contrast between highly myelinated and less myelinated brain regions. The T1w/T2w ratio is particularly useful because it can be derived from standard clinical MRI sequences and requires fairly minimal postprocessing, making it an ideal option for clinical translation. Previous studies show that T1w/T2w maps capture established patterns of cortical myelination, with higher ratios in sensory and motor regions and lower ratios in association cortices (5,6). Furthermore, T1w/T2w ratios have been used often in studies of multiple sclerosis as an index of demyelination and to describe lesions (7,8).

Neurodevelopmental disorders including autism and attention-deficit/-hyperactivity disorder (ADHD) are among those with suspected myelin abnormalities. However, studies of these conditions are often impeded by heterogeneity in etiology. This heterogeneity can be reduced through the adoption of a genetics-first approach. Therefore, in this study we examined disorders caused by single germline mutations in the RAS-extracellular signal-regulated mitogen-activated protein kinase (RAS-MAPK) signaling pathway. These conditions, collectively known as RASopathies, offer a unique opportunity to study neurobiological mechanisms within a genetically defined population. Furthermore, we can leverage preclinical models to inform our hypotheses and findings. Prior preclinical work indicated that causal mutations of RASopathies upregulate the RAS-MAPK pathway and lead to downstream cellular effects including disrupted oligodendrocyte maturation and fewer myelinated axons (9). Recent genetic evidence demonstrates that the RAS-MAPK pathway is likely involved in a variety of neurodevelopmental disorders with idiopathic etiologies such as autism and ADHD (10), suggesting that by studying RASopathies, we may also shed light on the brain pathology of broader neurodevelopmental disorders. Indeed, studies of youth with RASopathies have found elevated autism and ADHD symptoms (11). Through this genetics-first approach, we aimed to investigate the impact of genetic variants on cortical myelin patterns in children with RASopathies compared to typically developing (TD) peers, with potential relevance to a variety of neurodevelopmental disorders.

Prior studies in pediatric samples consistently show the expected spatial patterns in T1w/T2w ratios, with higher values in primary sensory regions compared to higher-order association areas (12,13). However, associations between T1w/T2w ratios and cognitive or behavioral measures are inconsistent. For example, a study of 621 youth aged 3-21 years from the Pediatric Imaging, Neurocognition, and Genetics (PING) study showed lower cortical T1w/T2w ratios were associated with better cognitive performance (12). Conversely, a study of 960 children aged 8-11 years selected at random from the Adolescent Brain Cognitive Development (ABCD) study found no significant relationships with cognitive measures (13). In neurodevelopmental conditions, results are similarly mixed. Some studies in children with autism report no group differences in cortical T1w/T2w ratios (14), while others have found widespread reductions and negative correlations between T1w/T2w and autism symptom severity based on the Childhood Autism Rating Scales (15). Additionally, a recent study of children with autism, ADHD, and TD peers reported no case-control differences in T1w/T2w ratios, though multimodal clustering indicated distinct cortical profiles that transcended diagnostic boundaries (16). The heterogeneity of these findings across studies reinforce the need for a genetics-first approach to studying neurodevelopmental mechanisms in order to uncover more specific patterns of brain organization.

In this study, we investigated T1w/T2w ratios as a proxy for cortical myelin content in youth with RASopathies compared to TD peers. Based on preclinical evidence of disrupted oligodendrocyte maturation (17), we hypothesized that youth with RASopathies would show reduced cortical T1w/T2w ratios, reflecting lower intracortical myelin content. Given that the T1w/T2w signal is more robust in cortex than in white matter (18,19), we focused our analysis on the cortical grey matter. The cerebral cortex is a convoluted structure composed of six architectonic layers, in which basic neuroscience studies have shown a gradient of decreasing myelination from the grey-white boundary to the cortical surface (20). While our image resolution (1 mm isotropic) is not high enough to distinctly resolve these separate layers, others have demonstrated layer-specific analysis, particularly using high-field MRI (21–23). Inspired by this knowledge, we explored the T1w/T2w ratio at the grey/white boundary, midline, and cortical surface. As a secondary exploratory aim, we examined whether cortical T1w/T2w ratios were associated with cognition and parent-rated physical health. This analysis was motivated by mixed findings in prior work, and the lack of studies specifically examining these relationships in RASopathies.

## Methods and Materials

### Participants

Participants were recruited from January 2023 through April 2025. Participants with RASopathies were recruited across the United States and Canada, while TD participants were recruited from the San Francisco Bay Area. Written informed consent was provided by legal guardians and complementary written assent was provided by participants aged over 7 years. All procedures in this work involving human subjects were approved by the Stanford University School of Medicine Institutional Review Board. Sub-groups of these participants were previously reported in prior works (24–27), however, this is the first study to report on the T1w/T2w ratio data.

The NIH Toolbox Cognition battery (Version 2) was administered to participants (28). The yielded scores were the Fluid (i.e., information processing, speed, and learning), Crystallized (i.e., fund of knowledge), and Total (i.e., Fluid and Crystallized) Composites, all of which yield T-scores (M=50, SD=10), with higher scores reflecting stronger cognitive abilities.

Additionally, the Mobility and Strength Impact portions from the Patient-Reported Outcomes Measurement Information System (PROMIS) were administered to parents (29,30). The Mobility portion asks the parent to report on the child’s daily gross motor functioning in the past seven days (e.g., ability to run, ride a bike, walk upstairs unassisted). The Strength Impact portion asks the parent to report on the child’s daily physical strength/stamina in the past seven days (e.g., strong enough to hold things above their head, jump up and down, open a jar by themselves). Both portions yield T-scores (M=50, SD=10), with higher scores indicating better health.

### Imaging Protocol

Prior to the MRI, participants completed behavioral training in a mock scanner to reduce motion and anxiety during the scan. MRI data were acquired using a standard 48-channel head coil on a GE Premier 3.0 Tesla whole-body system (GE Healthcare, Milwaukee, WI). A whole-brain high-resolution T1-weighted magnetization-prepared rapid gradient-echo (MPRAGE) sequence was used to acquire structural T1 data with the following parameters: repetition time (TR)= 1992 ms; echo time (TE)= 2.8 ms; inversion time (TI)= 900 ms; flip angle=8°; matrix size= 240 × 240; slice thickness= 1.2 mm; acquisition time (TA)=4 minutes 24 seconds. A 3D isotropic fast-spin echo sequence (CUBE) was used to acquire structural T2 data: TR= 3000 ms; TE= 100 ms; matrix size= 240 × 240; 1 mm isotropic voxel size; TA=4 minutes 58 seconds.

### Image Analysis

We ran the FreeSurfer (version 7.2, http://surfer.nmr.mgh.harvard.edu) default pipeline with the 3T flag. All outputs were examined and, when necessary, manual editing was conducted by author MR in accordance with FreeSurfer tutorial guidelines and re-run through the pipeline until acceptable. The FreeSurfer pipeline generated a bias-corrected T1w image (nu.mgz), and this was used as the T1w reference image (heretofore referred to simply as the T1w image) for the analysis. The T2w image was registered to the T1w image using *mri_coreg* (FreeSurfer), and intensity-normalized to the same scale using *mri_convert* with the resample-like (rl) flag. Bias-correction was conducted using the ANTS software package (31). The T1w/T2w ratio maps were created by dividing the corrected T1w volume by the T2w volume; example maps are shown in **Figure S1**. All ratio maps were subsequently quality checked by author JP.

To assess the myelination gradient across the cortex, we took a similar approach to a prior T1w/T2w ratio study in healthy aging adults (21). We chose to examine the cortex at three levels: at the cortical surface, grey/white boundary, and halfway between (midline). In order to minimize partial volume effects, we sampled the cortical surface at 0.5 mm below the FreeSurfer-defined pial surface. Likewise, we sampled the grey/white boundary 0.5 mm above the FreeSurfer-defined white surface (**see Figure 1**). The importance of this approach for minimizing partial volume effects is highlighted by **Figure S2**. We then used *mri_vol2surf* to average the T1w/T2w ratio values for each of the 68 Desikan-Killiany parcellated cortical regions-of-interest (ROIs) at each of these three levels (32,33). Additionally, average T1w/T2w ratios were extracted from subcortical ROIs by creating a mask for each region and subsequently eroding the label mask by 1 mm with a 2D kernel. Eight subcortical ROIs were investigated: accumbens, amygdala, caudate, hippocampus, pallidum, putamen, thalamus, and the ventral diencephalon.

**Figure 1.**
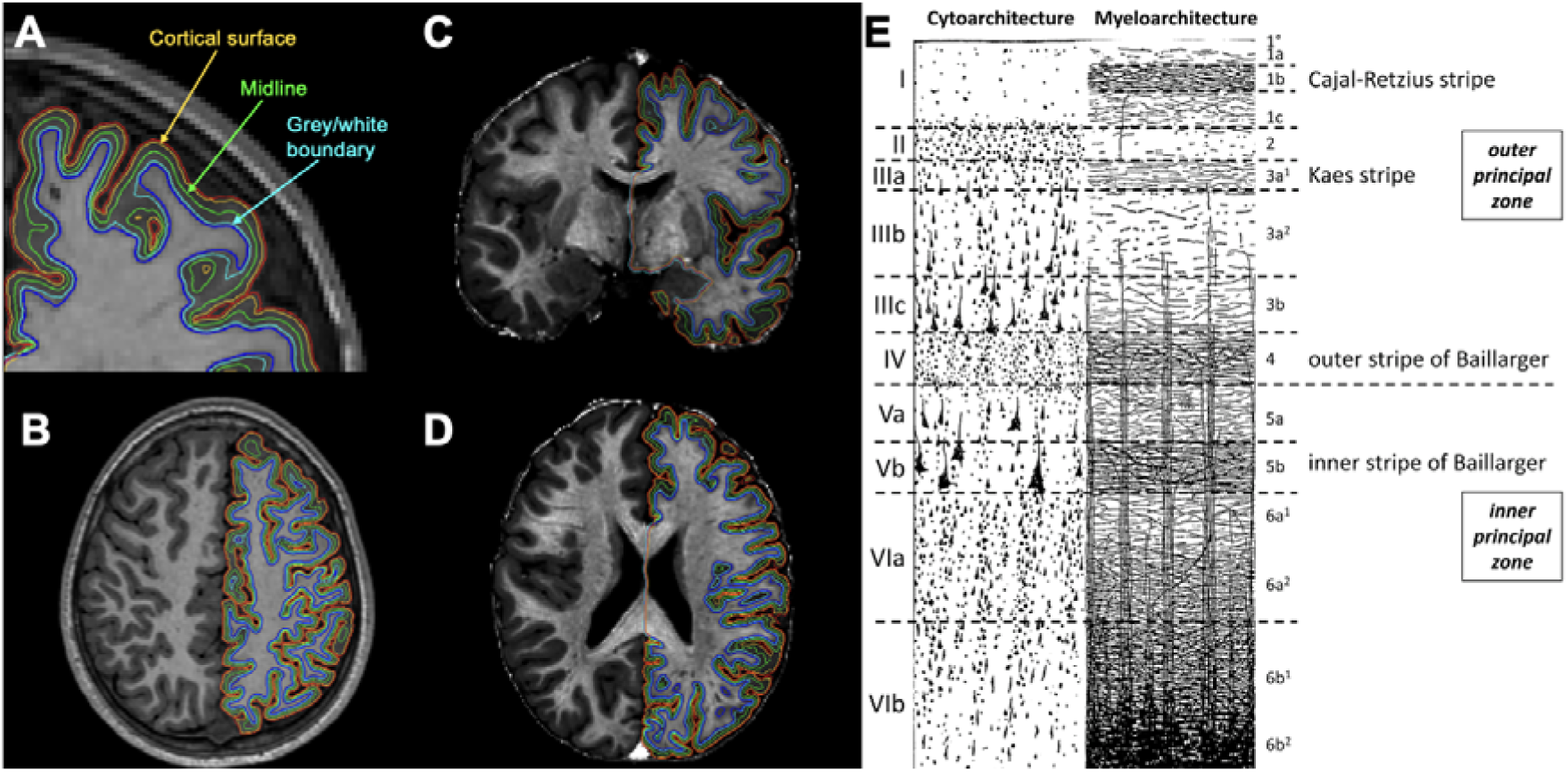
A) Coronal B) axial T1-weighted images C) coronal and D) axial T1w/T2w ratio images, showing the three levels at which the cortex was sampled: at the cortical surface (yellow line, 0.5 mm below the pial surface red line), midline (green line) and grey/white boundary (turquoise line, 0.5 mm above the white matter surface dark blue line); E) shows the cytoarchitecture and myeloarchitecture of the cortical layers, reproduced from Palomero-Gallagher and Zilles (2019) (1)

### Statistical Analysis

For cortical T1w/T2w ratio analysis, we used an analysis of covariance to assess between-groups differences (RASopathies, TD) in the average T1w/T2w ratio at each level (cortical surface, midline, grey/white boundary) for each of the 68 cortical regions, including age, sex, cortical thickness, curvature, and Euler number (as an index of motion) as covariates to control for potential confounding effects. For example, cortical thickness and curvature may be associated with myelin content (21,34). The false discovery rate (FDR) was used to correct for multiple comparisons. The same methods were applied to the between-groups analysis of eight subcortical regions, though cortical thickness and curvature were not included as covariates

For our exploratory aim, to reduce multiple comparisons, we first examined relationships between behavioral outcomes with an average whole-brain T2w/T2w ratio. If significant results were found, we investigated further using average T1w/T2w ratios from six cortical regions aggregated from the Desikan-Killiany atlas as follows: cingulate, frontal, insula, parietal, occipital, and temporal (see **Table S1** for groupings). We focused on the midline only for this brain-behavior analyses given the exploratory nature of this aim, and because the midline is least likely to incur partial volume effects. To assess if there was a relationship between T1w/T2w and the behavioral measures, we first conducted correlations. Pearson correlations were used except in cases of non-normal residuals, in which case Spearman correlations were used. Correlations were conducted separately in each group (RASopathies, TD) and across the entire sample. A Fisher’s r-to-z transformation was used to test for differences between the slopes in RASopathies versus TD.

To further probe these relationships, we performed group comparisons using a quartile-based approach. The average T1w/T2w value across the brain was calculated for each subject, and these T1w/T2 ratio values were then used to assign each participant to one of four quartiles. Quartile 1 represented the lowest T1w/T2w ratio values (indicating lower myelination) whereas Quartile 4 indicated the highest T1w/T2w ratio values (indicating higher myelination). We then compared the behavioral scores between each quartile using an ANCOVA with age, sex and RASopathy status included as factors. If appropriate, post-hoc Tukey’s pairwise t-tests were conducted. This quartile-based approach was chosen in place of a data-driven clustering method. Prior work used spectral-based clustering of the T1w/T2w ratio (16); however, our sample was too small to yield stable and generalizable clusters. Thus, this approach enabled us to test whether variation in cortical myelination was related to behavioral outcomes, within the constraints of our modest sample size.

## Results

### Participants

A total of 112 subjects (M_age_=10.7, SD_age_=3.38, age range 6.05 – 17.8 years; 54 female, 58 male) were included in the analysis. Of these subjects, 86 were in the RASopathies group (age range 6.05 – 17.8 years) and 26 were in the TD group (age range 6.13 – 16.8 years). There were no differences in age, sex, or motion (indexed by Euler number) between the two groups. **Table 1** shows the demographic and descriptive statistics. The genetic variants included in the RASopathies group are described in detail in the **Supplement**. Still, most are youth with the *PTPN11* genetic variant causing Noonan syndrome (n=34) or the *NF1* variant causing neurofibromatosis type 1 (NF1, n=18).

**Table 1.** Demographics and descriptive statistics of included participants.

|  | **TD (n=26)** | **RAS (n=86)** | **$t/\chi^2$** | **$p$** |
| --- | --- | --- | --- | --- |
| **Age** | 10.4 (3.38) | 10.8 (3.39) | 0.55 | .586 |
| **Sex (M/F)** | 12/14 | 44/42 | 0.00 | .987 |
| **Euler number** | -76.5 (30.5) | -63.8 (32.6) | 1.83 | .074 |
| **Fluid reasoning** | 48.8 (11.3) | 34.4 (8.74) | 5.97 | <.001 |
| **Crystallized intelligence** | 51.5 (12.0) | 39.9 (7.74) | 4.64 | <.001 |
| **Total cognition** | 50.1 (11.6) | 34.0 (7.99) | 6.56 | <.001 |
| **Mobility** | 53.0 (5.85) | 44.4 (5.82) | 6.55 | <.001 |
| **Strength impact** | 47.3 (7.52) | 42.0 (7.18) | 3.19 | .003 |

Data presented as counts or mean (SD). \*Three participants from the RASopathies group was excluded from cognitive measures due to missing data (RASopathies n=83). Fluid reasoning, crystallized intelligence, and total cognition are composite T-scores from the NIH Toolbox. Mobility and strength impact are T-scores derived from PROMIS ratings.

### Average T1w/T2w ratios show similar anatomical patterns across the brain in RASopathies and TD

Previous T1w/T2w ratio studies identified a consistent pattern of signal distribution, where sensorimotor regions show greater signal compared to the frontal lobe and temporal pole (5). A similar pattern is observed in cortical maps from our subjects with RASopathies and TD (**Figure 2**).

**Figure 2.**
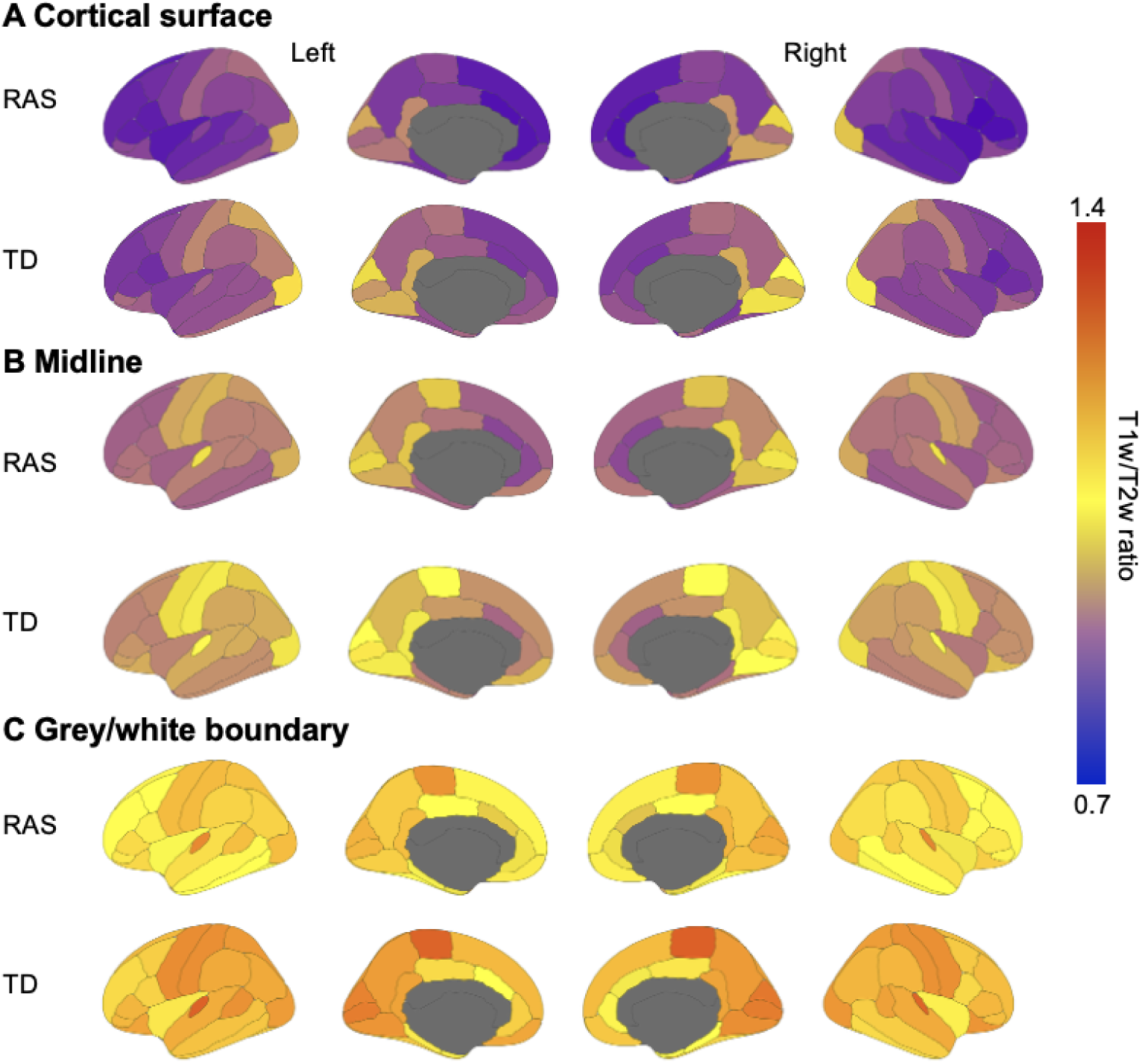
Cortical T1w/T2w ratio maps in RASopathies and TD sampled from A) the cortical surface B) the midline, and C) grey/white boundary. Colder colors indicate lower T1w/T2w ratios and warmer colours indicate greater T1w/T2w ratios. The maps for TD appear warmer (i.e., greater T1w/T2w ratio) than NS in A, B, and C. The overall signal distribution across the whole brain in NS and TD suggest higher T1w/T2w in sensorimotor regions, consistent with prior work. *RAS=RASopathies; TD=typical developing; T1w=T1-weighted; T2w=T2-weighted*.

### T1w/T2w ratio is significantly reduced in majority of cortical regions in RASopathies

Analysis of 68 cortical regions revealed widespread reductions in the T1w/T2w ratio in RASopathies compared to TD. Thus, although the broad spatial patterns are similar, the subjects with RASopathies show noticeably lower T1w/T2w signal across the brain compared to TD.

Of the T1w/T2w ratios sampled along the cortical surface, 50 regions (out of 68 total) were significantly reduced in RASopathies (*p_FDR_*<.050; **Table S2**).The largest effect sizes (where Cohen’s *d*>0.75) were in the left rostral anterior cingulate (*d*=1.01, *p_FDR_*<.001), left lateral occipital (*d*=0.980, *p_FDR_*=.001), right temporal pole (*d*=0.850, *p_FDR_*=.001), right rostral anterior cingulate (*d*=0.775, *p_FDR_*=.006), left medial orbitofrontal (d=0.774, *p_FDR_*=.006), and right superior temporal (*d*=0.769, *p_FDR_*=.006) regions.

Of the T1w/T2w ratios sampled from the midline, 63 regions of 68 were significantly reduced in RASopathies (*p_FDR_*<.050; **Table S3**). **Figures 3A and 3B** show the average T1w/T2w ratios in the left and right hemisphere regions, respectively. The largest effect sizes were in the left rostral anterior cingulate (*d*=0.777, *p_FDR_*=.002) and left pericalcarine (*d*=0.751, *p_FDR_*<.001). **Figure S3** shows the distribution of average T1w/T2w ratio values in selected cortical regions in RASopathies compared to TD.

**Figure 3.**
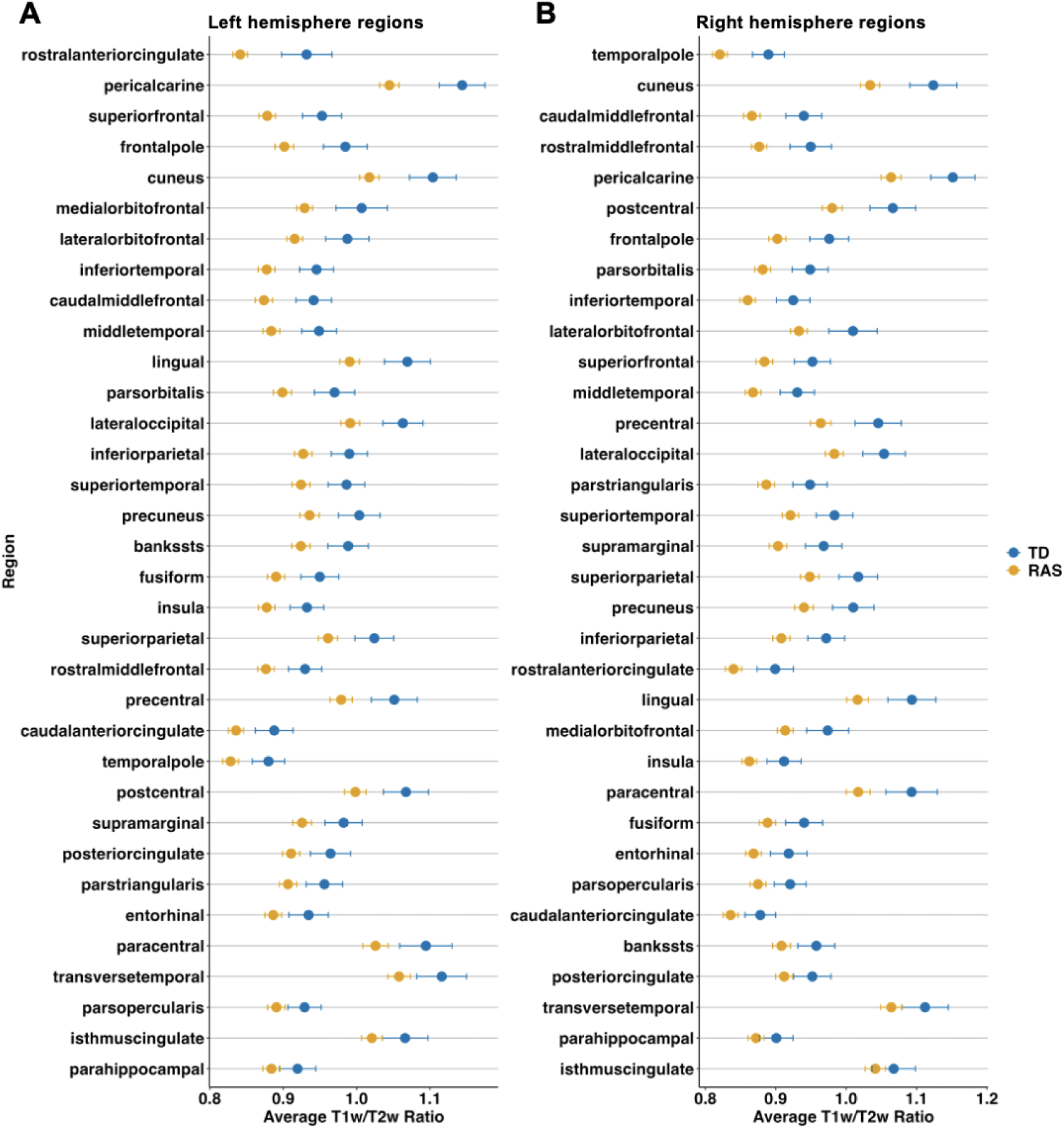
Average T1w/T2w ratios and standard errors for each cortical region in descending order from largest to smallest effect size: A) left hemisphere regions and B) right hemisphere regions. T1w/T2w ratios shown were sampled from midline. Effect sizes calculated using Cohen’s d. *RAS=RASopathies; TD=typical developing; T1w=T1-weighted; T2w=T2-weighted*.

Of the T1w/T2w ratios sampled from the grey/white matter boundary, 60 of 68 regions were significantly reduced in RASopathies compared to TD (*p_FDR_*<.050; **Table S4**). The largest effect sizes were in the left rostral anterior cingulate (*d*=0.791, *p_FDR_*=.003) and the right temporal pole (*d*=0.767, *p_FDR_*=.003). **Figures 4A, 4B, and 4C** shows the effect sizes projected onto the cortical regions for below grey matter, midline, above white matter samplings, respectively.

**Figure 4.**
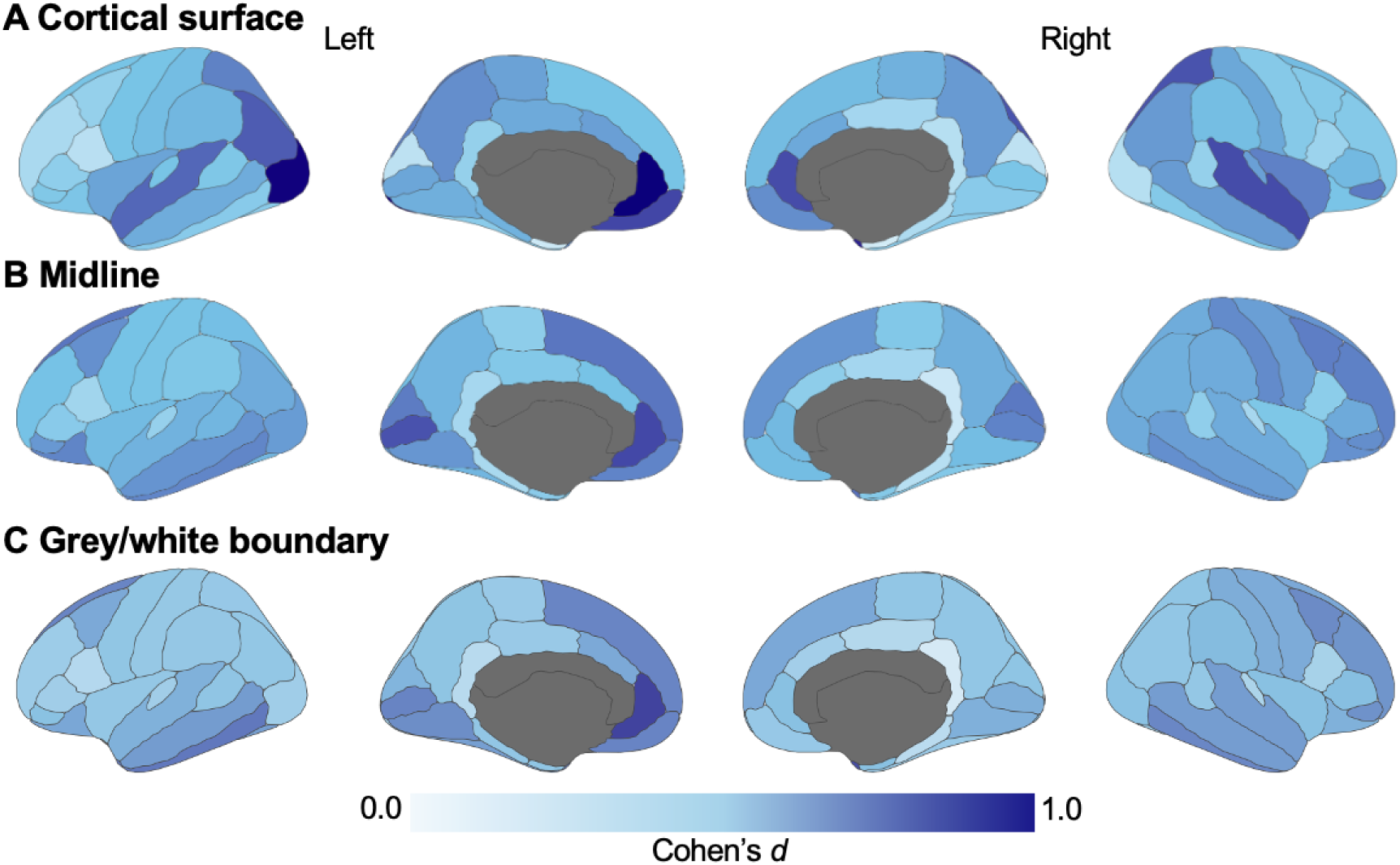
Effect size maps of the cortical regions from comparisons of T1w/T2w between RASopathies and TD. Effect sizes are shown as absolute values of Cohen’s *d*. All T1w/T2w values were higher in TD compared to RASopathies. Figure created using *ggseg* package in R v.4.4.1. *TD=typical developing; T1w=T1-weighted; T2w=T2-weighted*.

We did not find any evidence of sex differences in T1w/T2w ratios within each group (**Figure S4**). Furthermore, the pattern of reduced T1w/T2w ratio in RASopathies compared to TD appears fairly consistent within age bands (**Figure S5**). There were no significant differences in cortical T1w/T2w ratios between Noonan syndrome and NF1 groups (**Figure S6**).

### The cortical T1w/T2w ratio is, on average, 6% higher in TD compared to RASopathies

Given the large number of regions with reduced T1w/T2w ratios in RASopathies compared to TD, we additionally investigated whether the difference can be explained by a uniform scaling factor. To estimate the scaling factor, we fit three linear regression models – one each for the cortical surface, midline, and grey/white matter boundary regions. In all three models, we found strong linear relationships. The models based on regional T1w/T2w from the cortical surface and from the midline both estimated slopes of 1.069 (*R^2^*=0.99, *p*<.001, **Figures S7A, S7B**). The model based on T1w/T2w from the grey/white matter boundary fit a slope of 1.064 (*R^2^*=0.99, *p*<.001, **Figure S7C**). These results suggest the T1w/T2w values in the TD group are, on average, 6.4 to 6.9% higher than those in the RASopathies group.

### T1w/T2w ratio is significantly reduced in the accumbens in RASopathies but not in other subcortical regions

Analysis of T1w/T2w ratios of the subcortical regions revealed a significant reduction in T1w/T2w ratio in the RASopathies group (M=0.976, SD=0.120) relative to TD (M=1.046, SD=0.179), *d*=0.520, *p_FDR_*=.024. Initially, reductions in T1w/T2w ratios were also observed in the amygdala, caudate, and hippocampus, however these results did not survive FDR correction. The results in all subcortical regions investigated are described in **Table 2**. **Figure 5A** projects the effect sizes onto each subcortical region, and **Figure 5B** shows the distribution of average T1w/T2w values in the accumbens.

**Table 2.**
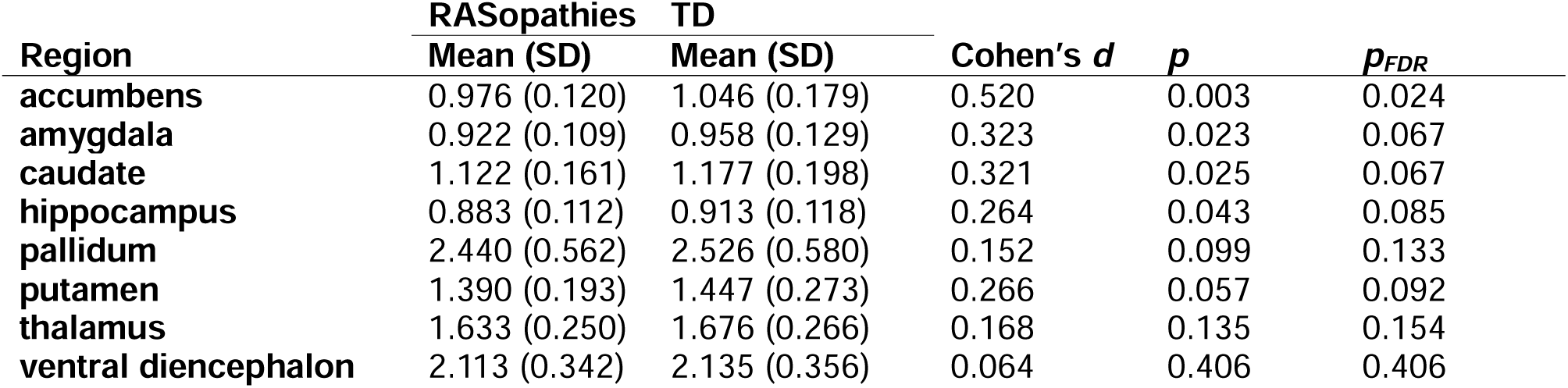
Comparison of T1w/T2w ratios in subcortical regions between RASopathies and TD.

**Figure 5.**
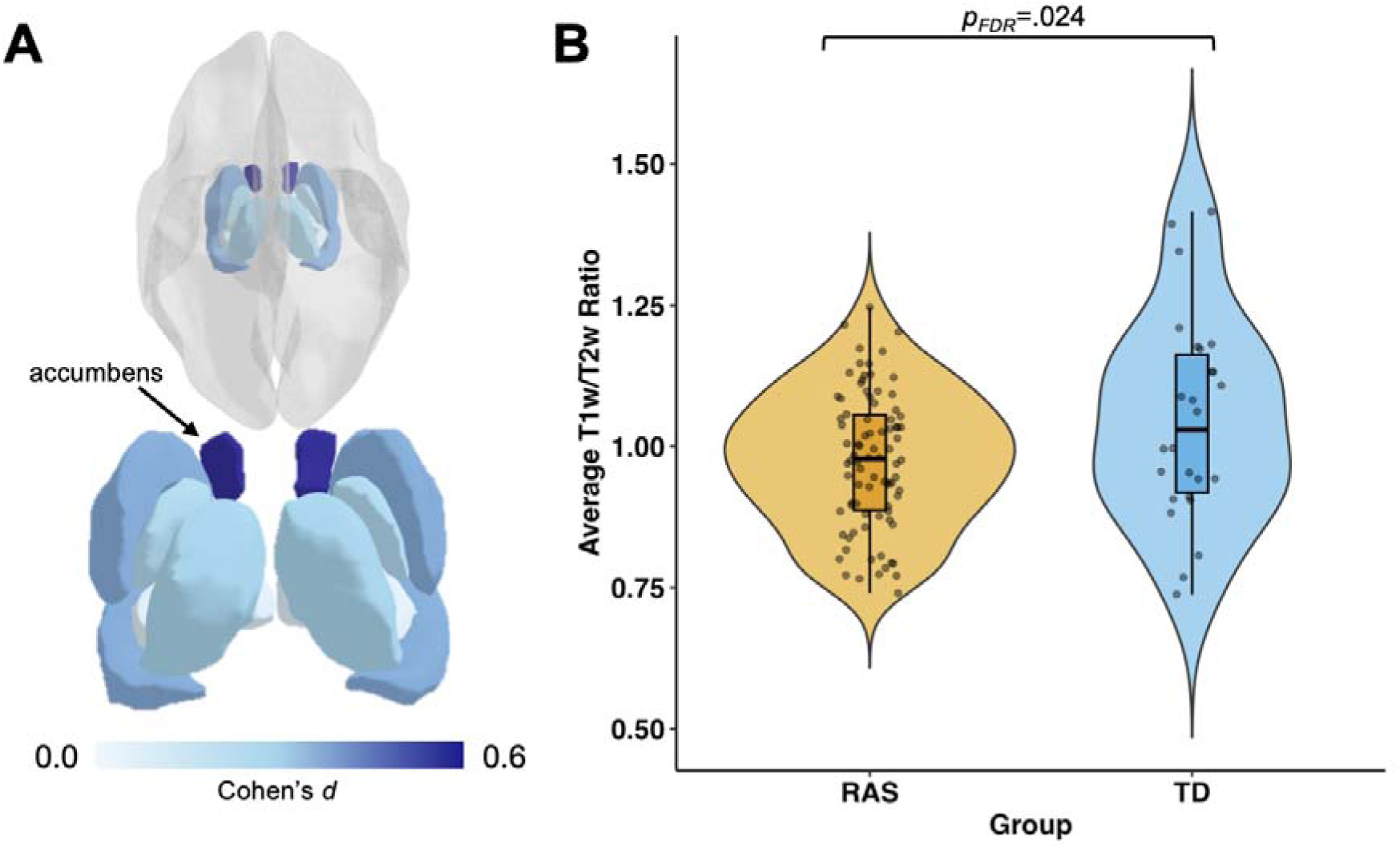
T1w/T2w ratio comparisons between RASopathies and TD in subcortical regions: A) Cohen’s *d* effect sizes show the greatest effect in the accumbens (dark blue); B) a violin plot of the distribution of average T1w/T2w ratio values in the accumbens in each group. All subcortical T1w/T2w values were higher in TD compared to RASopathies; however, the accumbens was the only subcortical region to show a significant difference between-groups (*p_FDR_*=.024). Cohen’s *d* values shown are the absolute values. Figure created using *ggseg* and *ggplot2* packages in R v.4.4.1. *RAS=RASopathies; TD=typical developing; T1w=T1-weighted; T2w=T2-weighted*.

### Mobility and strength are associated with cortical T1w/T2w ratios

We tested associations between the average whole-brain cortical T1w/T2w ratio and physical health and cognitive outcomes. Analyses were first conducted separately within each group (RASopathies, TD); however, Fisher’s r-to-z transformation indicated no significant differences between the slopes (**Tables S6-8**), so correlations were subsequently examined in the combined sample. The whole-brain T1w/T2w ratio was significantly correlated with mobility ratings (*r_s_*=.226, *p*=.017), strength impact ratings (*r_s_*=.310, *p*=.001), and weakly with fluid reasoning scores (*r*=0.195, *p*=.042); but not with crystallized intelligence (*r*=0.075, *p*=.438), nor total cognition (*r*=0.161, *p*=.094; **Figure S8**). To test if certain regions may be driving the associations, we further examined correlations between each outcome and an average T1w/T2w ratio from six aggregated cortical regions (i.e., cingulate, frontal, insula, occipital, parietal, and temporal). Significant Spearman correlations were found between all six regions and both PROMIS mobility and strength impact ratings (*p_FDR_*<.05; **Table S9),** however none of the correlations with cognitive composites survived FDR correction **(Table S10).**

To further test whether variation in cortical myelination was related to behavioral outcomes, we grouped each participant into one of four quartiles according to their average T1w/T2w ratio across the cortex (i.e., the first quartile has the lowest T1w/T2w ratio, while the fourth quartile has the highest T1w/T2w ratio). This analysis revealed significant effects of T1w/T2w ratio quartile (*p=*.002) and RASopathy status (*p<*.001) on mobility ratings. Post-hoc Tukey’s Tests showed significant differences in PROMIS Mobility ratings between youth in the first and third (*p*=.016) and the first and fourth (*p*=.004) T1w/T2w ratio quartiles (**Figure 6A**). We also found significant effects of the T1w/T2w ratio quartile (*p<*.001) and RASopathy status (*p=*.003) on PROMIS Strength Impact ratings. Post-hoc Tukey’s Tests revealed significant differences in PROMIS Strength ratings between youth in the first and fourth (*p*=.013), second and third (*p*=.043), and second and fourth (*p*=.010) T1w/T2w ratio quartiles (**Figure 6B**). There were no significant effects of the whole-brain T1w/T2w ratio quartile on the cognitive composite scores (**Figure S9**).

**Figure 6.**
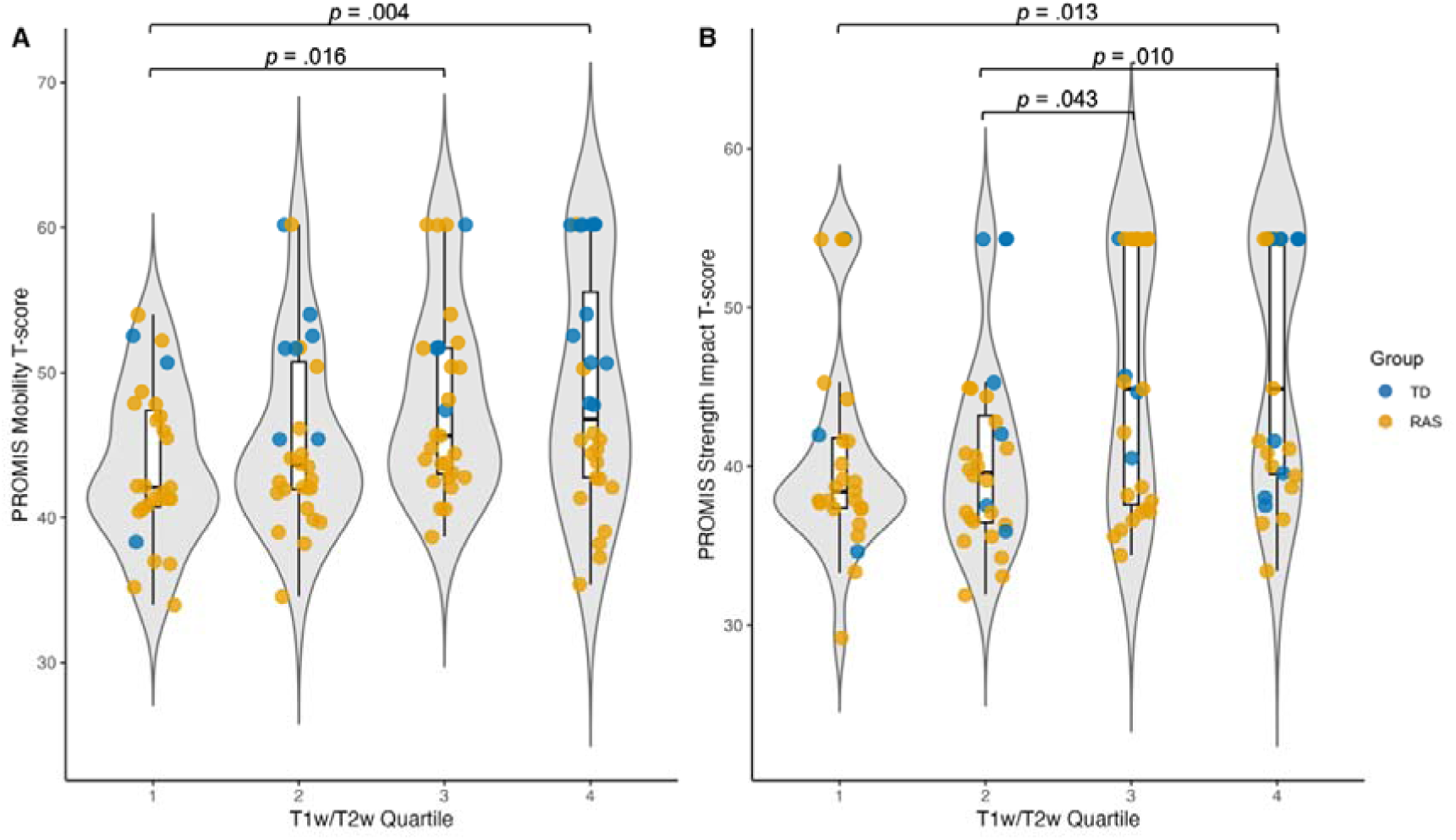
Differences between whole-brain T1w/T2w ratio quartiles in A) PROMIS Mobility ratings, and B) PROMIS Strength Impact ratings, with datapoints color-coded by group. The p-values shown are not corrected for multiple comparisons, given the exploratory nature of this aim. *RAS=RASopathies; TD=typical developing; T1w=T1-weighted; T2w=T2-weighted.* 1. Palomero-Gallagher N, Zilles K. Cortical layers: Cyto-, myelo-, receptor- and synaptic architecture in human cortical areas. Neuroimage. 2019 Aug 15;197:716–41.

Analyses of the six cortical regions were explored to examine whether any specific region was driving the significant mobility and strength impact results; however, the pattern of results was the same as the whole brain analysis (**Table S11-12**). There were no significant T1w/T2w ratio quartiles x RASopathy status interaction effects in the above analyses, thus, the interaction terms were removed from the final models presented above. However, a Chi-Square analysis revealed a significant difference in the proportions of RASopathy and TD participants across quartiles, Χ^2^(3)=9.81, *p*=.020. The composition of each quartile is shown in **Table 3**.

**Table 3.** Summary statistics of whole-brain T1w/T2w ratio by quartile.

| **Quartile** | **T1w/T2w ratio** | **Mobility** | **Strength** | **Fluid** | **Crystal** | **Total** | **RASopathies** | **TD** |
| --- | --- | --- | --- | --- | --- | --- | --- | --- |
|  | **Mean (SD)** | **Mean (SD)** | **Mean (SD)** | **Mean (SD)** | **Mean (SD)** | **Mean (SD)** | **n (%)** | **n (%)** |
| **1** | 0.788 (0.046) | 43.5 (5.34) | 40.6 (6.62) | 35.5 (11.0) | 40.7 (9.43) | 35.2 (10.5) | 25 (89) | 3 (11) |
| **2** | 0.895 (0.026) | 45.4 (6.34) | 40.5 (6.03) | 37.5 (9.22) | 45.7 (9.35) | 39.5 (9.82) | 21 (75) | 7 (25) |
| **3** | 0.983 (0.025) | 48.0 (6.46) | 45.5 (7.96) | 36.6 (9.21) | 40.3 (8.94) | 35.6 (9.13) | 24 (86) | 4 (14) |
| **4** | 1.08 (0.056) | 48.7 (8.05) | 46.5 (7.91) | 41.5 (14.2) | 44.1 (12.1) | 41.0 (14.3) | 16 (57) | 12 (43) |

*Mobility and Strength ratings are from the PROMIS Mobility and PROMIS Strength Impact portions, respectively. Fluid, crystal, and total refer to the cognition composite scales from the NIH Toolbox. The presented T1w/T2w ratio is the average across the entire cortical midline.*

## Discussion

This study provides evidence of widespread reductions in T1w/T2w ratio across cortical regions in children and adolescents with RASopathies compared to TD peers, suggesting reduced cortical myelin content. Although the overall spatial pattern of T1w/T2w distribution was similar between-groups – with sensorimotor regions showing the highest signal – youth with RASopathies exhibited lower signal intensity across nearly all sampled cortical regions. The accumbens was the only subcortical region to show a significant T1w/T2w reduction. Furthermore, cortical T1w/T2w ratios interacted with group to significantly predict VCI and FSIQ. T1w/T2w ratios were positively associated with VCI and FSIQ in TD youth, but not in RASopathies, suggesting a disrupted relationship between cortical myelin and cognition in this group. These findings provide preliminary evidence that T1w/T2w-derived microstructural metrics may serve as candidate biomarkers for altered cortical development and cognition in RASopathies.

While few studies have examined T1w/T2w ratios in children with RASopathies, a recent preprint showed similar reductions in T1w/T2w across the cortical lobes of children with neurofibromatosis type 1 (NF1) (35). Eighteen participants in our sample carried an *NF1* gene mutation, which disrupts neurofibromin function and leads to overactivation of the RAS-MAPK pathway (36). Oligodendrocytes, myelin-producing glial cells, contain neurofibromin and preclinical evidence implicates the *NF1* mutation in oligodendrocyte development and myelin regulation. Animal models of *NF1* show abnormal myelin phenotypes such as white matter decompaction, hypermyelination, and increased myelin thickness (37,38), while diffusion tensor imaging studies in humans with NF1 also report reductions in white matter structural connectivity (25,39–41). Our prior work using quantitative T1 mapping found elevated white matter myelin in children with RASopathies (27), yet the current findings suggest a contrasting profile in cortical grey matter. Although fewer studies have addressed cortical myelin, histological and imaging data in NF1 point to decreased cortical lamination (42) and reduced corticocortical connectivity (43). Supporting this regional dissociation, prior work in zebrafish models of NF1 found increased proliferation of neuroglial progenitor cells in the brainstem but not in the cortex (44). Postmortem analyses also reveal neuronal heterotopias in white matter, suggesting that neurons fail to properly migrate from their origin in the white matter to their intended destinations in the cortex (42). This disrupted migration may reflect a broader shift in neuron and oligodendrocyte density, with increased cellular presence in white matter and reduced density in cortical regions. While we cannot directly test this hypothesis here, our findings highlight the need for further investigation into the tissue-specific effects of *NF1* on myelin development.

Alongside youth with NF1, our sample included a large number of youth with Noonan syndrome. Noonan syndrome is a RASopathy caused predominantly by genetic variants encoding the SHP2 protein (e.g., *PTPN11, SOS1, RAF1*), ultimately leading to hyperactivation of the RAS-MAPK pathway. Animal models of Noonan syndrome caused by *PTPN11* suggest reductions in astrocyte formation and axon myelination, along with promotion of neurogenesis (45,46). Imaging studies have also reported decreases in structural connectivity, similar to those observed in NF1 (25,47). Although there are distinctions between NF1 and Noonan syndrome in molecular mechanisms and neurodevelopmental phenotypes, our findings suggest similar reductions in cortical T1w/T2w ratios across both Noonan syndrome and NF1, with no significant differences observed between the groups. These results suggest potential convergence on disrupted cortical myelination within the RAS-MAPK pathway. A limitation of the current study is that we grouped individuals with different genetic variants into the single RASopathies category. Although patterns were broadly similar across variant groups in our sample, future studies with larger cohorts may be able to explore whether specific genetic mutations yield distinguishable neurobiological profiles.

Results from our exploratory aim revealed a significant effect of the T1w/T2w ratio on gross motor and stamina/strength across participants. Specifically, youth in the first or second quartiles of the T1w/T2w ratio (i.e., lower ratios, thus lower myelin content) tended to have worse health outcomes than those in the third and fourth quartiles (i.e., higher ratios, thus higher myelin content). Although a quartile x RASopathy status interaction was not found, this is likely due to the few TD participants in the lower quartiles. In other words, while there was no significant interaction effect, given that participants with RASopathies tended to be in the lower quartiles (especially compared to TD participants), this finding is particularly relevant for youth with RASopathies. Indeed, myelin content via the T1w/T2w ratio may be an emerging biomarker of motor and stamina abilities of youth with RASopathies. This is important given that motor difficulties are prevalent in youth with RASopathies, and gross motor abilities in particular worsen relative to normative data and unaffected siblings over time (48). Interestingly, unlike prior research (11, 44) our findings did not yield strong relationships between cognition and the T1w/T2w ratio. Importantly, the literature is mixed; earlier studies in TD youth have reported positive, negative, and absent associations depending on age, brain region, and cognitive domain (12,13,49,50).

While T1w/T2w is used widely as a proxy for intracortical myelin, the signal is influenced by other biological properties including water and iron content and does not provide a direct measure of myelin (51–54). The reliability of the signal has also been questioned in white matter, prompting us to focus on cortical regions where the signal is perhaps more robust (18,55). Although T1w/T2w offers a clinically practical approach, techniques like myelin water imaging may be more specific for quantifying myelin content (19). Additional limitations include our broad age range (6-17 years) and the use of parent-rated health outcomes that may not capture objective measures of functional impairments. Future work may consider investigating the T1w/T2w ratio and health outcomes within narrower age bands in youth with RASopathies to determine if the associations between cortical myelin content and physical health persist.

To our knowledge, this is the first work to probe intracortical and subcortical myelin content using the T1w/T2w ratio technique in a group of youth with RASopathies, including those with genetic variants linked to both NF1 and Noonan syndrome. Our findings reveal widespread reductions in cortical myelin in this group, potentially reflecting underlying disruptions in neuronal migration and oligodendrocyte development observed in preclinical models. Furthermore, the association between cortical myelin and motor functioning and stamina may point to the relevance of the T1w/T2w ratio as a biomarker for these difficulties. These results provide new insights into cortical brain development in RASopathies and highlight the need for future work to determine their clinical significance and potential for informing targeted interventions.

## Data Availability

All data produced in the present study are available upon reasonable request to the authors.

## Acknowledgments

We thank the families who participated in this research. The authors would also like to thank the Noonan Syndrome Foundation and the RASopathies Network which made this work possible. We would like to thank Stanford University and the Stanford Research Computing Center for providing computational resources and support that contributed to these research results, some of the computing for this project was performed on the Sherlock cluster. We gratefully acknowledge the support of The Lucas Service Center at Stanford.

