## Supplementary Material for "T1w/T2w ratio suggests reduced intracortical myelin content in youth with RASopathies"

**Methods and Materials**

**Participants**

*Inclusion criteria: RASopathies*

Proof of genetic testing was required for individuals with RASopathies to demonstrate the presence of appropriate mutations. The inclusion criteria for all participants with RASopathies are as follows: a) age from 5 years, 0 months to 17 years, 11 months; and b) gestational age > 34 weeks. Participants with neurofibromatosis type 1 and focal areas of signal intensity with no mass effect and/or low-grade optic pathway glioma are not excluded.

*Inclusion criteria: typical developing (TD)*

TD participants are group-matched to participants with RASopathies based on age, sex, handedness, socioeconomic status, pubertal status, and ethnicity. Subjects are included if their full-scale IQ is greater than 80.

*Exclusion criteria*

For all participants, the exclusion criteria includes: a) presence of neurological or psychiatric disease (e.g., psychotic symptoms); b) sensory deficits that would preclude participation in assessments or imaging; c) history of significant head trauma with loss of consciousness; d) use of psychotropic medications or currently taking medications with central nervous system effects within 5 half-lives of the scan; e) contraindications to MRI (e.g., metal implants, orthodontia, claustrophobia); f) history of alcohol or drug use; g) premature birth (<34 weeks); h) low birth weight (<2000g); i) history of head trauma with loss of consciousness; neurological disorders known to affect cognitive development or brain structure; j) known presence of gliomas in cerebellum, brainstem, or basal ganglia; and k) contraindications to MRI.

Additionally, participants with neurofibromatosis type 1 were excluded if they received an MEK inhibitor or chemotherapy, and/or show presence of cerebellar, brainstem, tectal plate, and basal ganglia gliomas. Participants with RASopathies were not excluded if they show clinical symptoms associated with typical syndromic cognitive-behavioral features (e.g., inattention, anxiety, learning disorder, or autistic traits).

Pubertal status was determined using Tanner staging (1,2). Full-scale, performance, and verbal IQs were acquired using the Wechsler Abbreviated Scale of Intelligence 2^nd^ Edition (3).

**Imaging Protocol**

Participants completed behavioral training in a mock scanner prior to the actual MRI in order to reduce anxiety and motion during the MRI. During the MRI, to minimize motion artifacts and anxiety, participants were provided with a video to watch of their choice. Additionally, scanner personnel used an MRI-compatible camera affixed to the head coil focused on the participant’s face to check for motion in real-time during the scan. Scans were stopped and restarted if excessive motion was observed during the scan. After each scan acquisition, images were checked for artifacts by the scan operator and scans were repeated in cases of poor quality. At the beginning of the MRI and in-between scan acquisitions, participants were reminded to stay as still as possible.

**Sensitivity Analysis**

T1-weighted and T2-weighted structural images were visually inspected by a trained radiologist for the presence of T2-hyperintensities. To test if T2-hyperintensities might drive between-group differences in subcortical regions, a sensitivity analysis was performed excluding all subjects with identified T2-hyperintensities.

**Results**

**Power calculation and sample size justification**

Given the absence of prior work using T1w/T2w ratios in RASopathies, there are no previous studies on which to base our sample size justification. We collected a final sample for analysis of 86 subjects with RASopathies and 26 TD. At an α=0.05 and 1-β=0.8, the minimum detectable effect size is therefore *d*=0.663.

**Participants**

The following genetic variants were reported among included participants with RASopathies: *PTPN11* (n=34), *NF1* (n=18) *SOS1* (n=11), *RAF1* (n=5), Noonan syndrome with multiple lentigines (NSML) (n=4), *NRAS* (n=3), *RIT1* (n=3), *SOS2* (n=3), *KRAS* (n=2), *LZTR1* (n=2), and *SHOC2* (n=1). Of the participants with an *NF1* genetic variant, one of these participants reported with NF1 microdeletion syndrome (otherwise known as 17q11.2 deletion syndrome) indicated by a 1.4-Mb loss on chromosome 17 at q11.2. Of the participants with NSML, all four reported with *PTPN11* heterozygous genetic variants, with amino acid substitutions at p.Thr468Met (n=3) and p.Tyr279Cys (n=1). NSML is a condition distinct from *PTPN11* genetic variants causing NS, e.g., gain-of-function *PTPN11* variants cause NS, whereas loss-of-function variants cause NSML. However, both NS and NSML are disorders of the RAS-MAPK pathway and therefore grouped under the RASopathies umbrella. For more information on the genetics underlying NS and NSML, see (4,5), and for information on NF1 see (6,7).

**Sensitivity Analysis**

Twenty-five subjects showed the presence of T2-hyperintensities (*NF1*=14, *PTPN11*=9, *RAF1*=2). After excluding the subjects with T2-hyperintensities from analyses of the subcortical regions, there were no significant differences between-groups. The T1w/T2w ratio in the accumbens initially showed a difference between RASopathies and TD; however, the finding did not remain significant following FDR correction. The results of the sensitivity analysis are shown in **Table S5**.

**Table S1. Groupings of Desikan-Killiany ROIs into larger regions.**

| **Larger region** | **Desikan-Killiany ROIs** |
| --- | --- |
| Frontal | caudalmiddlefrontal, rostralmiddlefrontal, superiorfrontal, parsopercularis, parsorbitalis, parstriangularis, lateralorbitofrontal, medialorbitofrontal, precentral, frontalpole |
| Temporal | superiortemporal, middletemporal, inferiortemporal, fusiform, temporalpole, transversetemporal, entorhinal, parahippocampal |
| Parietal | inferiorparietal, superiorparietal, supramarginal, postcentral, precuneus |
| Occipital | lateraloccipital, lingual, cuneus, pericalcarine |
| Cingulate | rostralanteriorcingulate, caudalanteriorcingulate |
| Insula | insula |

**Table S2. Cortical region comparisons (TD v RASopathies) of T1w/T2w ratios sampled from the cortical surface.**

|  |  | **TD** | | **RASopathies** | |  |  |  |
| --- | --- | --- | --- | --- | --- | --- | --- | --- |
| **Hemi** | **Region** | **Mean** | **SE** | **Mean** | **SE** | ***p*** | ***p_FDR_*** | ***d*** |
| LH | caudalmiddlefrontal | 0.832 | 0.019 | 0.796 | 0.011 | 0.083 | 0.092 | 0.358 |
| LH | entorhinal | 0.888 | 0.026 | 0.865 | 0.014 | 0.041 | 0.055 | 0.173 |
| LH | postcentral | 0.937 | 0.029 | 0.879 | 0.012 | 0.014 | **0.024** | 0.470 |
| LH | parstriangularis | 0.812 | 0.018 | 0.775 | 0.009 | 0.022 | **0.033** | 0.418 |
| LH | supramarginal | 0.884 | 0.021 | 0.837 | 0.010 | 0.018 | **0.030** | 0.492 |
| LH | insula | 0.817 | 0.022 | 0.765 | 0.010 | 0.008 | **0.017** | 0.560 |
| LH | bankssts | 0.857 | 0.020 | 0.811 | 0.010 | 0.004 | **0.012** | 0.484 |
| LH | lateralorbitofrontal | 0.874 | 0.021 | 0.832 | 0.010 | 0.068 | 0.081 | 0.442 |
| LH | parsorbitalis | 0.892 | 0.028 | 0.837 | 0.013 | 0.027 | **0.037** | 0.442 |
| LH | middletemporal | 0.852 | 0.019 | 0.802 | 0.010 | 0.020 | **0.032** | 0.546 |
| LH | pericalcarine | 0.957 | 0.025 | 0.888 | 0.012 | <.001 | **0.002** | 0.612 |
| LH | parahippocampal | 0.852 | 0.023 | 0.797 | 0.011 | 0.002 | **0.006** | 0.526 |
| LH | paracentral | 0.906 | 0.026 | 0.842 | 0.012 | 0.007 | **0.016** | 0.550 |
| LH | medialorbitofrontal | 0.874 | 0.024 | 0.803 | 0.009 | 0.002 | **0.006** | 0.774 |
| LH | frontalpole | 0.887 | 0.039 | 0.819 | 0.016 | 0.020 | **0.032** | 0.418 |
| LH | cuneus | 1.051 | 0.045 | 0.985 | 0.022 | 0.080 | 0.091 | 0.317 |
| LH | inferiortemporal | 0.909 | 0.022 | 0.856 | 0.013 | 0.090 | 0.099 | 0.447 |
| LH | rostralmiddlefrontal | 0.811 | 0.017 | 0.781 | 0.009 | 0.116 | 0.123 | 0.355 |
| LH | rostralanteriorcingulate | 0.851 | 0.024 | 0.759 | 0.009 | <.001 | **<.001** | 1.009 |
| LH | isthmuscingulate | 0.987 | 0.026 | 0.940 | 0.012 | 0.026 | **0.037** | 0.393 |
| LH | lateraloccipital | 1.156 | 0.045 | 0.992 | 0.016 | <.001 | **0.001** | 0.980 |
| LH | lingual | 0.995 | 0.036 | 0.914 | 0.014 | 0.001 | **0.005** | 0.579 |
| LH | superiorparietal | 0.982 | 0.039 | 0.896 | 0.013 | 0.006 | **0.014** | 0.611 |
| LH | parsopercularis | 0.795 | 0.017 | 0.772 | 0.009 | 0.139 | 0.141 | 0.270 |
| LH | fusiform | 0.854 | 0.023 | 0.808 | 0.011 | 0.008 | **0.018** | 0.437 |
| LH | caudalanteriorcingulate | 0.808 | 0.020 | 0.755 | 0.009 | 0.003 | **0.009** | 0.588 |
| LH | superiorfrontal | 0.812 | 0.017 | 0.770 | 0.009 | 0.007 | **0.015** | 0.494 |
| LH | temporalpole | 0.776 | 0.021 | 0.719 | 0.010 | 0.003 | **0.010** | 0.586 |
| LH | precuneus | 0.877 | 0.020 | 0.814 | 0.010 | 0.000 | **0.004** | 0.657 |
| LH | transversetemporal | 0.885 | 0.023 | 0.828 | 0.012 | 0.013 | **0.023** | 0.493 |
| LH | precentral | 0.863 | 0.022 | 0.814 | 0.010 | 0.027 | **0.037** | 0.497 |
| LH | inferiorparietal | 0.928 | 0.031 | 0.843 | 0.011 | 0.001 | **0.006** | 0.706 |
| LH | posteriorcingulate | 0.871 | 0.021 | 0.819 | 0.010 | 0.001 | **0.006** | 0.555 |
| LH | superiortemporal | 0.849 | 0.019 | 0.784 | 0.010 | 0.001 | **0.006** | 0.724 |
| RH | caudalmiddlefrontal | 0.823 | 0.017 | 0.782 | 0.011 | 0.125 | 0.129 | 0.416 |
| RH | entorhinal | 0.899 | 0.027 | 0.874 | 0.016 | 0.061 | 0.074 | 0.166 |
| RH | postcentral | 0.919 | 0.025 | 0.859 | 0.012 | 0.010 | **0.021** | 0.533 |
| RH | parstriangularis | 0.797 | 0.017 | 0.753 | 0.010 | 0.005 | **0.014** | 0.497 |
| RH | supramarginal | 0.856 | 0.021 | 0.810 | 0.011 | 0.027 | **0.037** | 0.460 |
| RH | insula | 0.810 | 0.020 | 0.756 | 0.009 | 0.013 | **0.023** | 0.603 |
| RH | bankssts | 0.835 | 0.019 | 0.794 | 0.010 | 0.011 | **0.021** | 0.434 |
| RH | lateralorbitofrontal | 0.885 | 0.023 | 0.837 | 0.010 | 0.117 | 0.123 | 0.498 |
| RH | parsorbitalis | 0.857 | 0.024 | 0.792 | 0.011 | 0.002 | **0.008** | 0.615 |
| RH | middletemporal | 0.830 | 0.017 | 0.779 | 0.009 | 0.050 | 0.063 | 0.581 |
| RH | pericalcarine | 0.990 | 0.034 | 0.910 | 0.015 | 0.001 | **0.006** | 0.548 |
| RH | parahippocampal | 0.812 | 0.020 | 0.784 | 0.009 | 0.027 | **0.037** | 0.316 |
| RH | paracentral | 0.900 | 0.027 | 0.837 | 0.011 | 0.001 | **0.006** | 0.558 |
| RH | medialorbitofrontal | 0.857 | 0.024 | 0.797 | 0.009 | 0.021 | **0.032** | 0.619 |
| RH | frontalpole | 0.895 | 0.029 | 0.812 | 0.014 | 0.001 | **0.006** | 0.612 |
| RH | cuneus | 1.113 | 0.051 | 1.044 | 0.021 | 0.043 | 0.056 | 0.322 |
| RH | inferiortemporal | 0.879 | 0.020 | 0.836 | 0.011 | 0.145 | 0.145 | 0.407 |
| RH | rostralmiddlefrontal | 0.814 | 0.018 | 0.773 | 0.010 | 0.103 | 0.111 | 0.458 |
| RH | rostralanteriorcingulate | 0.832 | 0.022 | 0.755 | 0.010 | 0.001 | **0.006** | 0.775 |
| RH | isthmuscingulate | 0.987 | 0.027 | 0.958 | 0.013 | 0.069 | 0.081 | 0.245 |
| RH | lateraloccipital | 1.075 | 0.042 | 1.019 | 0.016 | 0.073 | 0.084 | 0.332 |
| RH | lingual | 1.059 | 0.040 | 0.979 | 0.017 | 0.003 | **0.010** | 0.468 |
| RH | superiorparietal | 0.975 | 0.039 | 0.878 | 0.012 | 0.000 | **0.004** | 0.715 |
| RH | parsopercularis | 0.781 | 0.017 | 0.749 | 0.009 | 0.054 | 0.067 | 0.375 |
| RH | fusiform | 0.842 | 0.022 | 0.798 | 0.010 | 0.004 | **0.012** | 0.452 |
| RH | caudalanteriorcingulate | 0.805 | 0.019 | 0.757 | 0.009 | 0.005 | **0.013** | 0.577 |
| RH | superiorfrontal | 0.817 | 0.016 | 0.773 | 0.010 | 0.011 | **0.021** | 0.505 |
| RH | temporalpole | 0.799 | 0.019 | 0.721 | 0.010 | <.001 | **0.001** | 0.850 |
| RH | precuneus | 0.883 | 0.022 | 0.817 | 0.010 | <.001 | **0.002** | 0.659 |
| RH | transversetemporal | 0.884 | 0.022 | 0.819 | 0.012 | 0.011 | **0.022** | 0.570 |
| RH | precentral | 0.839 | 0.021 | 0.796 | 0.011 | 0.014 | **0.024** | 0.434 |
| RH | inferiorparietal | 0.887 | 0.025 | 0.824 | 0.011 | 0.006 | **0.015** | 0.601 |
| RH | posteriorcingulate | 0.860 | 0.020 | 0.828 | 0.010 | 0.046 | 0.059 | 0.328 |
| RH | superiortemporal | 0.843 | 0.019 | 0.774 | 0.009 | 0.001 | **0.006** | 0.769 |

*d=absolute value of Cohen’s d; FDR=false discovery rate; hemi=hemisphere; LH=left hemisphere; RH=right hemisphere; SE=standard error*

**Table S3. Cortical region comparisons (TD v RASopathies) of T1w/T2w ratios sampled from the midline.**

|  | |  | **TD** | | **RASopathies** | |  |  |  |
| --- | --- | --- | --- | --- | --- | --- | --- | --- | --- |
| **Hemi** | **Region** | | **Mean** | **SE** | **Mean** | **SE** | ***p*** | ***p_FDR_*** | ***d*** |
| LH | caudalmiddlefrontal | | 0.942 | 0.024 | 0.874 | 0.012 | 0.002 | **0.005** | 0.603 |
| LH | entorhinal | | 0.935 | 0.027 | 0.886 | 0.011 | 0.007 | **0.013** | 0.426 |
| LH | postcentral | | 1.067 | 0.031 | 0.998 | 0.015 | 0.006 | **0.012** | 0.489 |
| LH | parstriangularis | | 0.956 | 0.025 | 0.906 | 0.012 | 0.018 | **0.024** | 0.434 |
| LH | supramarginal | | 0.982 | 0.025 | 0.926 | 0.013 | 0.003 | **0.006** | 0.469 |
| LH | insula | | 0.932 | 0.023 | 0.877 | 0.011 | 0.004 | **0.009** | 0.517 |
| LH | bankssts | | 0.988 | 0.027 | 0.924 | 0.013 | 0.002 | **0.005** | 0.526 |
| LH | lateralorbitofrontal | | 0.987 | 0.029 | 0.916 | 0.011 | 0.010 | **0.014** | 0.629 |
| LH | parsorbitalis | | 0.970 | 0.027 | 0.899 | 0.012 | 0.008 | **0.013** | 0.586 |
| LH | middletemporal | | 0.949 | 0.024 | 0.884 | 0.011 | 0.007 | **0.013** | 0.597 |
| LH | pericalcarine | | 1.144 | 0.031 | 1.045 | 0.013 | <.001 | **<.001** | 0.751 |
| LH | parahippocampal | | 0.920 | 0.025 | 0.884 | 0.012 | 0.006 | **0.011** | 0.317 |
| LH | paracentral | | 1.094 | 0.036 | 1.026 | 0.017 | 0.001 | **0.004** | 0.414 |
| LH | medialorbitofrontal | | 1.007 | 0.035 | 0.929 | 0.011 | 0.008 | **0.013** | 0.633 |
| LH | frontalpole | | 0.985 | 0.030 | 0.902 | 0.013 | 0.001 | **0.004** | 0.657 |
| LH | cuneus | | 1.104 | 0.032 | 1.017 | 0.013 | <.001 | **0.001** | 0.650 |
| LH | inferiortemporal | | 0.945 | 0.023 | 0.877 | 0.011 | 0.021 | **0.027** | 0.626 |
| LH | rostralmiddlefrontal | | 0.930 | 0.023 | 0.876 | 0.011 | 0.060 | 0.061 | 0.501 |
| LH | rostralanteriorcingulate | | 0.932 | 0.034 | 0.841 | 0.010 | <.001 | **0.002** | 0.777 |
| LH | isthmuscingulate | | 1.066 | 0.031 | 1.021 | 0.014 | 0.050 | 0.053 | 0.328 |
| LH | lateraloccipital | | 1.063 | 0.027 | 0.991 | 0.013 | 0.038 | **0.043** | 0.581 |
| LH | lingual | | 1.069 | 0.031 | 0.991 | 0.013 | <.001 | **0.002** | 0.591 |
| LH | superiorparietal | | 1.024 | 0.026 | 0.961 | 0.013 | 0.003 | **0.008** | 0.506 |
| LH | parsopercularis | | 0.929 | 0.023 | 0.891 | 0.012 | 0.048 | 0.051 | 0.346 |
| LH | fusiform | | 0.950 | 0.026 | 0.890 | 0.012 | 0.002 | **0.005** | 0.526 |
| LH | caudalanteriorcingulate | | 0.888 | 0.026 | 0.836 | 0.010 | 0.011 | **0.016** | 0.496 |
| LH | superiorfrontal | | 0.953 | 0.027 | 0.878 | 0.011 | <.001 | **0.001** | 0.665 |
| LH | temporalpole | | 0.880 | 0.022 | 0.828 | 0.011 | 0.007 | **0.013** | 0.493 |
| LH | precuneus | | 1.004 | 0.028 | 0.936 | 0.013 | 0.001 | **0.004** | 0.529 |
| LH | transversetemporal | | 1.116 | 0.034 | 1.058 | 0.015 | 0.010 | **0.014** | 0.387 |
| LH | precentral | | 1.051 | 0.031 | 0.979 | 0.015 | 0.010 | **0.014** | 0.500 |
| LH | inferiorparietal | | 0.990 | 0.025 | 0.927 | 0.012 | 0.001 | **0.005** | 0.548 |
| LH | posteriorcingulate | | 0.964 | 0.027 | 0.911 | 0.012 | 0.006 | **0.011** | 0.457 |
| LH | superiortemporal | | 0.986 | 0.025 | 0.924 | 0.012 | 0.008 | **0.013** | 0.533 |
| RH | caudalmiddlefrontal | | 0.940 | 0.025 | 0.866 | 0.012 | 0.004 | **0.008** | 0.643 |
| RH | entorhinal | | 0.918 | 0.026 | 0.868 | 0.011 | 0.006 | **0.012** | 0.448 |
| RH | postcentral | | 1.066 | 0.032 | 0.980 | 0.014 | 0.001 | **0.005** | 0.616 |
| RH | parstriangularis | | 0.949 | 0.024 | 0.887 | 0.012 | 0.001 | **0.004** | 0.551 |
| RH | supramarginal | | 0.968 | 0.026 | 0.903 | 0.012 | 0.001 | **0.004** | 0.547 |
| RH | insula | | 0.912 | 0.024 | 0.863 | 0.011 | 0.030 | **0.037** | 0.474 |
| RH | bankssts | | 0.958 | 0.026 | 0.908 | 0.013 | 0.010 | **0.014** | 0.407 |
| RH | lateralorbitofrontal | | 1.010 | 0.034 | 0.933 | 0.012 | 0.051 | 0.053 | 0.601 |
| RH | parsorbitalis | | 0.949 | 0.026 | 0.882 | 0.011 | 0.003 | **0.008** | 0.608 |
| RH | middletemporal | | 0.930 | 0.024 | 0.868 | 0.011 | 0.033 | **0.038** | 0.574 |
| RH | pericalcarine | | 1.152 | 0.031 | 1.064 | 0.014 | <.001 | **0.001** | 0.634 |
| RH | parahippocampal | | 0.901 | 0.024 | 0.872 | 0.011 | 0.034 | **0.039** | 0.264 |
| RH | paracentral | | 1.093 | 0.037 | 1.017 | 0.017 | 0.001 | **0.004** | 0.461 |
| RH | medialorbitofrontal | | 0.974 | 0.030 | 0.913 | 0.011 | 0.046 | **0.050** | 0.512 |
| RH | frontalpole | | 0.976 | 0.028 | 0.902 | 0.012 | <.001 | **0.003** | 0.612 |
| RH | cuneus | | 1.124 | 0.033 | 1.034 | 0.014 | <.001 | **<.001** | 0.656 |
| RH | inferiortemporal | | 0.925 | 0.024 | 0.860 | 0.011 | 0.009 | **0.013** | 0.606 |
| RH | rostralmiddlefrontal | | 0.950 | 0.029 | 0.876 | 0.011 | 0.020 | **0.026** | 0.639 |
| RH | rostralanteriorcingulate | | 0.899 | 0.026 | 0.840 | 0.012 | 0.033 | **0.038** | 0.519 |
| RH | isthmuscingulate | | 1.068 | 0.031 | 1.041 | 0.014 | 0.158 | 0.158 | 0.189 |
| RH | lateraloccipital | | 1.054 | 0.030 | 0.983 | 0.013 | 0.008 | **0.013** | 0.551 |
| RH | lingual | | 1.093 | 0.034 | 1.016 | 0.015 | <.001 | **0.003** | 0.513 |
| RH | superiorparietal | | 1.017 | 0.027 | 0.948 | 0.013 | 0.001 | **0.004** | 0.547 |
| RH | parsopercularis | | 0.920 | 0.023 | 0.875 | 0.011 | 0.016 | **0.022** | 0.422 |
| RH | fusiform | | 0.940 | 0.026 | 0.888 | 0.012 | 0.002 | **0.006** | 0.456 |
| RH | caudalanteriorcingulate | | 0.878 | 0.022 | 0.836 | 0.011 | 0.042 | **0.046** | 0.416 |
| RH | superiorfrontal | | 0.952 | 0.026 | 0.884 | 0.012 | 0.002 | **0.006** | 0.596 |
| RH | temporalpole | | 0.890 | 0.023 | 0.820 | 0.011 | <.001 | **0.003** | 0.677 |
| RH | precuneus | | 1.010 | 0.029 | 0.940 | 0.013 | 0.001 | **0.004** | 0.541 |
| RH | transversetemporal | | 1.112 | 0.033 | 1.064 | 0.015 | 0.046 | 0.050 | 0.324 |
| RH | precentral | | 1.046 | 0.033 | 0.964 | 0.015 | <.001 | **0.003** | 0.573 |
| RH | inferiorparietal | | 0.972 | 0.026 | 0.908 | 0.012 | 0.002 | **0.005** | 0.540 |
| RH | posteriorcingulate | | 0.952 | 0.027 | 0.912 | 0.012 | 0.023 | **0.028** | 0.337 |
| RH | superiortemporal | | 0.983 | 0.026 | 0.921 | 0.012 | 0.006 | **0.011** | 0.548 |

*d=absolute value of Cohen’s d; FDR=false discovery rate; hemi=hemisphere; LH=left hemisphere; RH=right hemisphere; SE=standard error*

**Table S4. Cortical region comparisons (TD v RASopathies) of T1w/T2w ratios sampled from the grey/white matter boundary.**

|  | |  | **TD** | | **RASopathies** | |  |  |  |
| --- | --- | --- | --- | --- | --- | --- | --- | --- | --- |
| **Hemi** | **Region** | | **Mean** | **SE** | **Mean** | **SE** | ***p*** | ***p_FDR_*** | ***d*** |
| LH | caudalmiddlefrontal | | 1.182 | 0.030 | 1.107 | 0.014 | 0.004 | **0.009** | 0.548 |
| LH | entorhinal | | 1.130 | 0.033 | 1.061 | 0.015 | 0.001 | **0.005** | 0.468 |
| LH | postcentral | | 1.285 | 0.034 | 1.213 | 0.017 | 0.012 | **0.018** | 0.442 |
| LH | parstriangularis | | 1.221 | 0.031 | 1.164 | 0.015 | 0.021 | **0.027** | 0.393 |
| LH | supramarginal | | 1.242 | 0.032 | 1.175 | 0.016 | 0.003 | **0.009** | 0.447 |
| LH | insula | | 1.142 | 0.028 | 1.084 | 0.014 | 0.004 | **0.009** | 0.449 |
| LH | bankssts | | 1.279 | 0.035 | 1.207 | 0.016 | 0.004 | **0.009** | 0.467 |
| LH | lateralorbitofrontal | | 1.250 | 0.039 | 1.167 | 0.014 | 0.014 | **0.019** | 0.554 |
| LH | parsorbitalis | | 1.220 | 0.036 | 1.145 | 0.015 | 0.017 | **0.024** | 0.493 |
| LH | middletemporal | | 1.200 | 0.030 | 1.120 | 0.014 | 0.004 | **0.009** | 0.576 |
| LH | pericalcarine | | 1.315 | 0.032 | 1.224 | 0.015 | <.001 | **0.003** | 0.629 |
| LH | parahippocampal | | 1.139 | 0.030 | 1.076 | 0.015 | 0.006 | **0.010** | 0.457 |
| LH | paracentral | | 1.359 | 0.042 | 1.277 | 0.020 | 0.003 | **0.009** | 0.427 |
| LH | medialorbitofrontal | | 1.245 | 0.054 | 1.138 | 0.013 | 0.006 | **0.010** | 0.628 |
| LH | frontalpole | | 1.207 | 0.044 | 1.115 | 0.015 | 0.002 | **0.006** | 0.557 |
| LH | cuneus | | 1.306 | 0.035 | 1.227 | 0.015 | <.001 | **0.003** | 0.523 |
| LH | inferiortemporal | | 1.195 | 0.030 | 1.107 | 0.014 | 0.008 | **0.013** | 0.657 |
| LH | rostralmiddlefrontal | | 1.165 | 0.030 | 1.110 | 0.014 | 0.110 | 0.113 | 0.406 |
| LH | rostralanteriorcingulate | | 1.180 | 0.049 | 1.057 | 0.012 | <.001 | **0.003** | 0.791 |
| LH | isthmuscingulate | | 1.252 | 0.035 | 1.206 | 0.016 | 0.093 | 0.098 | 0.293 |
| LH | lateraloccipital | | 1.264 | 0.028 | 1.209 | 0.015 | 0.188 | 0.191 | 0.399 |
| LH | lingual | | 1.276 | 0.036 | 1.181 | 0.016 | <.001 | **0.003** | 0.608 |
| LH | superiorparietal | | 1.269 | 0.031 | 1.203 | 0.016 | 0.018 | **0.024** | 0.438 |
| LH | parsopercularis | | 1.177 | 0.028 | 1.134 | 0.015 | 0.059 | 0.064 | 0.312 |
| LH | fusiform | | 1.197 | 0.033 | 1.116 | 0.014 | 0.001 | **0.004** | 0.569 |
| LH | caudalanteriorcingulate | | 1.102 | 0.033 | 1.030 | 0.013 | 0.004 | **0.009** | 0.547 |
| LH | superiorfrontal | | 1.216 | 0.040 | 1.122 | 0.014 | <.001 | **0.003** | 0.626 |
| LH | temporalpole | | 1.071 | 0.030 | 0.983 | 0.014 | <.001 | **0.003** | 0.644 |
| LH | precuneus | | 1.261 | 0.035 | 1.187 | 0.016 | 0.003 | **0.009** | 0.474 |
| LH | transversetemporal | | 1.364 | 0.040 | 1.295 | 0.018 | 0.006 | **0.010** | 0.393 |
| LH | precentral | | 1.290 | 0.036 | 1.211 | 0.018 | 0.022 | **0.027** | 0.464 |
| LH | inferiorparietal | | 1.241 | 0.030 | 1.177 | 0.015 | 0.005 | **0.009** | 0.451 |
| LH | posteriorcingulate | | 1.165 | 0.032 | 1.104 | 0.015 | 0.012 | **0.018** | 0.426 |
| LH | superiortemporal | | 1.234 | 0.032 | 1.156 | 0.015 | 0.004 | **0.009** | 0.531 |
| RH | caudalmiddlefrontal | | 1.191 | 0.034 | 1.103 | 0.014 | 0.005 | **0.009** | 0.613 |
| RH | entorhinal | | 1.114 | 0.030 | 1.044 | 0.015 | 0.001 | **0.004** | 0.497 |
| RH | postcentral | | 1.280 | 0.036 | 1.193 | 0.017 | 0.004 | **0.009** | 0.542 |
| RH | parstriangularis | | 1.219 | 0.033 | 1.145 | 0.015 | 0.002 | **0.006** | 0.504 |
| RH | supramarginal | | 1.221 | 0.032 | 1.145 | 0.015 | 0.002 | **0.006** | 0.511 |
| RH | insula | | 1.129 | 0.033 | 1.063 | 0.013 | 0.013 | **0.019** | 0.498 |
| RH | bankssts | | 1.255 | 0.035 | 1.185 | 0.016 | 0.005 | **0.009** | 0.448 |
| RH | lateralorbitofrontal | | 1.277 | 0.052 | 1.183 | 0.015 | 0.076 | 0.081 | 0.531 |
| RH | parsorbitalis | | 1.208 | 0.034 | 1.124 | 0.014 | 0.002 | **0.007** | 0.600 |
| RH | middletemporal | | 1.184 | 0.032 | 1.104 | 0.014 | 0.020 | **0.026** | 0.574 |
| RH | pericalcarine | | 1.317 | 0.032 | 1.236 | 0.016 | 0.001 | **0.004** | 0.536 |
| RH | parahippocampal | | 1.107 | 0.027 | 1.068 | 0.015 | 0.047 | 0.053 | 0.286 |
| RH | paracentral | | 1.365 | 0.041 | 1.279 | 0.020 | 0.004 | 0.009 | 0.456 |
| RH | medialorbitofrontal | | 1.193 | 0.034 | 1.130 | 0.014 | 0.074 | 0.080 | 0.437 |
| RH | frontalpole | | 1.193 | 0.040 | 1.111 | 0.015 | 0.001 | **0.004** | 0.516 |
| RH | cuneus | | 1.325 | 0.034 | 1.250 | 0.017 | <.001 | **0.003** | 0.472 |
| RH | inferiortemporal | | 1.165 | 0.031 | 1.082 | 0.014 | 0.005 | **0.009** | 0.621 |
| RH | rostralmiddlefrontal | | 1.209 | 0.047 | 1.116 | 0.013 | 0.022 | **0.027** | 0.593 |
| RH | rostralanteriorcingulate | | 1.124 | 0.033 | 1.054 | 0.014 | 0.043 | **0.048** | 0.496 |
| RH | isthmuscingulate | | 1.251 | 0.035 | 1.224 | 0.016 | 0.260 | 0.260 | 0.173 |
| RH | lateraloccipital | | 1.273 | 0.032 | 1.205 | 0.014 | 0.013 | **0.019** | 0.486 |
| RH | lingual | | 1.286 | 0.037 | 1.200 | 0.016 | 0.001 | **0.004** | 0.533 |
| RH | superiorparietal | | 1.259 | 0.032 | 1.188 | 0.016 | 0.005 | **0.009** | 0.466 |
| RH | parsopercularis | | 1.166 | 0.030 | 1.116 | 0.014 | 0.029 | **0.033** | 0.366 |
| RH | fusiform | | 1.175 | 0.033 | 1.113 | 0.015 | 0.003 | **0.009** | 0.427 |
| RH | caudalanteriorcingulate | | 1.068 | 0.026 | 1.019 | 0.013 | 0.029 | **0.034** | 0.383 |
| RH | superiorfrontal | | 1.212 | 0.037 | 1.130 | 0.015 | 0.005 | **0.009** | 0.549 |
| RH | temporalpole | | 1.074 | 0.031 | 0.968 | 0.014 | <.001 | **0.003** | 0.767 |
| RH | precuneus | | 1.266 | 0.035 | 1.193 | 0.017 | 0.006 | **0.010** | 0.458 |
| RH | transversetemporal | | 1.358 | 0.038 | 1.301 | 0.017 | 0.027 | **0.032** | 0.347 |
| RH | precentral | | 1.284 | 0.038 | 1.193 | 0.017 | 0.001 | **0.005** | 0.543 |
| RH | inferiorparietal | | 1.222 | 0.032 | 1.156 | 0.015 | 0.005 | **0.009** | 0.455 |
| RH | posteriorcingulate | | 1.146 | 0.030 | 1.101 | 0.015 | 0.025 | **0.030** | 0.316 |
| RH | superiortemporal | | 1.239 | 0.034 | 1.155 | 0.015 | 0.002 | **0.006** | 0.576 |

*d=absolute value of Cohen’s d; FDR=false discovery rate; hemi=hemisphere; LH=left hemisphere; RH=right hemisphere; SE=standard error*

**Table S5.** **Sensitivity analysis results: T1w/T2w ratios in subcortical regions following exclusion of subjects with T2-hyperintensities.**

| **Region** | **RASopathies: Mean (SD)** | **TD:**  **Mean (SD)** | ***p*** | ***p_FDR_*** | **Cohen’s *d*** |
| --- | --- | --- | --- | --- | --- |
| Accumbens | 0.976 (0.131) | 1.046 (0.179) | 0.013 | 0.102 | 0.478 |
| Amygdala | 0.930 (0.122) | 0.958 (0.129) | 0.123 | 0.242 | 0.231 |
| Caudate | 1.124 (0.177) | 1.177 (0.198) | 0.088 | 0.242 | 0.287 |
| Hippocampus | 0.887 (0.125) | 0.913 (0.118) | 0.151 | 0.242 | 0.208 |
| Pallidum | 2.487 (0.576) | 2.526 (0.580) | 0.450 | 0.600 | 0.067 |
| Putamen | 1.387 (0.214) | 1.447 (0.273) | 0.130 | 0.242 | 0.254 |
| Thalamus | 1.659 (0.271) | 1.676 (0.266) | 0.543 | 0.620 | 0.063 |
| Ventral diencephalon | 2.146 (0.368) | 2.135 (0.356) | 0.906 | 0.906 | -0.029 |

*Twenty-five subjects with T2-hyperintensities were excluded prior to analysis. The remaining subjects (total n=87) included RASopathies (n=61) and TD (n=26).*

**Table S6.** **Correlations between an average whole-brain T1w/T2w ratios and behavioral outcomes in each group.**

|  | **TD (n=26)** | | **RAS (n=84)** | |  |  |
| --- | --- | --- | --- | --- | --- | --- |
| **Measure** | ***r*** | ***p*** | ***r*** | ***p*** | **Fisher’s *z*** | ***p*** |
| Mobility | 0.332 | 0.098 | 0.089 | 0.420 | 1.083 | 0.279 |
| Strength impact | 0.319 | 0.112 | 0.242 | **0.028** | 0.354 | 0.723 |
| Crystallized | 0.086 | 0.677 | -0.108 | 0.330 | 0.824 | 0.410 |
| Fluid | 0.167 | 0.414 | 0.061 | 0.582 | 0.455 | 0.649 |
| Total | 0.158 | 0.440 | -0.026 | 0.819 | 0.784 | 0.433 |

Spearman correlations were conducted for correlations with mobility and strength impact, due to non-normal data. Pearson correlations were conducted for crystallized, fluid, and total cognition scores.

**Table S7.** **Spearman correlations between regional T1w/T2w ratios and PROMIS physical health outcomes in each group.**

|  |  | **TD** | | | **RAS** | | |  |  |  |
| --- | --- | --- | --- | --- | --- | --- | --- | --- | --- | --- |
| **Health measure** | **Region** | ***r*** | ***p*** | ***p_FDR_*** | ***r*** | ***p*** | ***p_FDR_*** | **Fisher’s *z*** | ***p*** | ***p_FDR_*** |
| Mobility | Cingulate | 0.337 | 0.092 | 0.110 | 0.088 | 0.423 | 0.678 | 1.115 | 0.265 | 0.266 |
|  | Frontal | 0.311 | 0.122 | 0.122 | 0.054 | 0.624 | 0.739 | 1.135 | 0.256 | 0.266 |
|  | Insula | 0.408 | **0.039** | 0.102 | 0.037 | 0.739 | 0.739 | 1.681 | 0.093 | 0.266 |
|  | Occipital | 0.386 | 0.051 | 0.102 | 0.130 | 0.237 | 0.678 | 1.177 | 0.239 | 0.266 |
|  | Parietal | 0.355 | 0.075 | 0.110 | 0.109 | 0.320 | 0.678 | 1.112 | 0.266 | 0.266 |
|  | Temporal | 0.393 | **0.047** | 0.102 | 0.083 | 0.452 | 0.678 | 1.409 | 0.159 | 0.266 |
| Strength impact | Cingulate | 0.387 | 0.051 | 0.138 | 0.227 | **0.039** | 0.058 | 0.753 | 0.451 | 0.901 |
|  | Frontal | 0.299 | 0.138 | 0.138 | 0.202 | 0.066 | 0.080 | 0.436 | 0.663 | 0.901 |
|  | Insula | 0.317 | 0.114 | 0.138 | 0.184 | 0.096 | 0.096 | 0.604 | 0.546 | 0.901 |
|  | Occipital | 0.330 | 0.100 | 0.138 | 0.279 | **0.011** | **0.038** | 0.241 | 0.810 | 0.901 |
|  | Parietal | 0.299 | 0.137 | 0.138 | 0.272 | **0.013** | **0.038** | 0.125 | 0.901 | 0.901 |
|  | Temporal | 0.363 | 0.069 | 0.138 | 0.231 | **0.036** | 0.058 | 0.614 | 0.539 | 0.901 |

*r_s_ is Spearman correlation r-value; p is uncorrected p-value; p_FDR_ is false discovery rate corrected p-value, corrected per health measure. Fisher’s r-to-z transformation was used to test differences between slopes.*

**Table S8.** **Peason correlations between regional T1w/T2w ratios and NIH Toolbox cognitive composite measures in each group.**

|  |  | **TD** | | | **RAS** | | |  |  |  |
| --- | --- | --- | --- | --- | --- | --- | --- | --- | --- | --- |
| **Cognitive measure** | **Region** | ***r*** | ***p*** | ***p_FDR_*** | ***r*** | ***p*** | ***p_FDR_*** | **Fisher’s *z*** | ***p*** | ***p_FDR_*** |
| Crystallized | Cingulate | 0.014 | 0.945 | 0.985 | -0.109 | 0.329 | 0.421 | 0.522 | 0.601 | 0.722 |
|  | Frontal | -0.004 | 0.985 | 0.985 | -0.087 | 0.432 | 0.432 | 0.356 | 0.722 | 0.722 |
|  | Insula | 0.074 | 0.719 | 0.985 | -0.135 | 0.223 | 0.421 | 0.893 | 0.372 | 0.558 |
|  | Occipital | 0.131 | 0.524 | 0.985 | -0.104 | 0.351 | 0.421 | 1.000 | 0.317 | 0.558 |
|  | Parietal | 0.136 | 0.508 | 0.985 | -0.122 | 0.273 | 0.421 | 1.100 | 0.272 | 0.558 |
|  | Temporal | 0.164 | 0.424 | 0.985 | -0.117 | 0.294 | 0.421 | 1.198 | 0.231 | 0.558 |
| Fluid | Cingulate | 0.089 | 0.665 | 0.665 | 0.030 | 0.787 | 0.845 | 0.251 | 0.802 | 0.802 |
|  | Frontal | 0.107 | 0.603 | 0.665 | 0.043 | 0.696 | 0.845 | 0.271 | 0.787 | 0.802 |
|  | Insula | 0.127 | 0.535 | 0.665 | 0.022 | 0.845 | 0.845 | 0.451 | 0.652 | 0.802 |
|  | Occipital | 0.196 | 0.337 | 0.665 | 0.055 | 0.624 | 0.845 | 0.610 | 0.542 | 0.802 |
|  | Parietal | 0.229 | 0.261 | 0.665 | 0.087 | 0.434 | 0.845 | 0.619 | 0.536 | 0.802 |
|  | Temporal | 0.208 | 0.307 | 0.665 | 0.069 | 0.536 | 0.845 | 0.605 | 0.545 | 0.802 |
| Total Cognition | Cingulate | 0.066 | 0.749 | 0.753 | -0.047 | 0.675 | 0.885 | 0.479 | 0.632 | 0.701 |
|  | Frontal | 0.065 | 0.753 | 0.753 | -0.025 | 0.819 | 0.885 | 0.384 | 0.701 | 0.701 |
|  | Insula | 0.127 | 0.537 | 0.753 | -0.068 | 0.543 | 0.885 | 0.829 | 0.407 | 0.611 |
|  | Occipital | 0.204 | 0.317 | 0.635 | -0.026 | 0.815 | 0.885 | 0.989 | 0.323 | 0.611 |
|  | Parietal | 0.227 | 0.265 | 0.635 | -0.016 | 0.885 | 0.885 | 1.049 | 0.294 | 0.611 |
|  | Temporal | 0.233 | 0.253 | 0.635 | -0.025 | 0.822 | 0.885 | 1.112 | 0.266 | 0.611 |

*r is Pearson correlation r-value; p is uncorrected p-value; p_FDR_ is false discovery rate corrected p-value, corrected per cognitive composite measure. Fisher’s r-to-z transformation was used to test differences between slopes.*

**Table S9.** **Spearman correlations between regional T1w/T2w ratios and PROMIS physical health outcomes.**

| **Health measure** | **Region** | ***r_s_*** | ***p*** | ***p_FDR_*** |
| --- | --- | --- | --- | --- |
| Strength impact | Cingulate | 0.297 | **0.002** | **0.003** |
|  | Frontal | 0.280 | **0.003** | **0.004** |
|  | Insula | 0.269 | **0.005** | **0.005** |
|  | Occipital | 0.344 | **<.001** | **0.001** |
|  | Parietal | 0.329 | **<.001** | **0.001** |
|  | Temporal | 0.297 | **0.002** | **0.003** |
| Mobility | Cingulate | 0.204 | **0.032** | **0.045** |
|  | Frontal | 0.192 | **0.043** | **0.045** |
|  | Insula | 0.190 | **0.045** | **0.045** |
|  | Occipital | 0.260 | **0.006** | **0.023** |
|  | Parietal | 0.252 | **0.008** | **0.023** |
|  | Temporal | 0.207 | **0.029** | **0.045** |

*r_s_ is Spearman correlation r-value; p is uncorrected p-value; p_FDR_ is false discovery rate corrected p-value, corrected per health measure.*

**Table S10.** **Peason correlations between regional T1w/T2w ratios and NIH Toolbox cognitive composite measures.**

| **Cognitive measure** | **Region** | ***r*** | ***p*** | ***p_FDR_*** |
| --- | --- | --- | --- | --- |
| Crystallized | Cingulate | 0.035 | 0.716 | 0.716 |
|  | Frontal | 0.069 | 0.474 | 0.711 |
|  | Insula | 0.049 | 0.612 | 0.716 |
|  | Occipital | 0.112 | 0.246 | 0.711 |
|  | Parietal | 0.081 | 0.402 | 0.711 |
|  | Temporal | 0.086 | 0.372 | 0.711 |
| Fluid | Cingulate | 0.140 | 0.145 | 0.145 |
|  | Frontal | 0.184 | 0.056 | 0.084 |
|  | Insula | 0.158 | 0.101 | 0.122 |
|  | Occipital | 0.223 | **0.020** | 0.059 |
|  | Parietal | 0.227 | **0.017** | 0.059 |
|  | Temporal | 0.205 | **0.033** | 0.065 |
| Total Cognition | Cingulate | 0.106 | 0.274 | 0.274 |
|  | Frontal | 0.151 | 0.117 | 0.175 |
|  | Insula | 0.124 | 0.198 | 0.237 |
|  | Occipital | 0.200 | **0.037** | 0.141 |
|  | Parietal | 0.185 | 0.055 | 0.141 |
|  | Temporal | 0.174 | 0.071 | 0.141 |

*r is Pearson correlation r-value; p is uncorrected p-value; p_FDR_ is false discovery rate corrected p-value, corrected per cognitive composite measure.*

**Table S11. Effects of T1w/T2w ratios in each region and RASopathy status on PROMIS Mobility ratings**

| Factor | *F* | *p* | Significant Quartile Differences* |
| --- | --- | --- | --- |
| Frontal Lobe | | | |
| T1w/T2w Ratio Quartile | 1.15 | .012 | 1^st^ & 3^rd^ (*p*=.034); 1^st^ and 4^th^ (*p*=.022) |
| Rasopathy Status | 7.89 | <.001 |  |
| Parietal Lobe | | | |
| T1w/T2w Ratio Quartile | 5.47 | .001 | 1^st^ and 4^th^ (*p*<.001) |
| Rasopathy Status | 34.97 | <.001 |  |
| Temporal Lobe | | | |
| T1w/T2w Ratio Quartile | 3.94 | .010 | 1^st^ and 4^th^ (*p*=.007) |
| Rasopathy Status | 40.10 | <.001 |  |
| Occipital Lobe | | | |
| T1w/T2w Ratio Quartile | 4.36 | .006 | 1^st^ & 3^rd^ (*p*=.028); 1^st^ and 4^th^ (*p*=.007) |
| Rasopathy Status | 39.71 | <.001 |  |
| Cingulate Cortex | | | |
| T1w/T2w Ratio Quartile | 4.44 | .005 | 1^st^ and 4^th^ (*p*=.002) |
| Rasopathy Status | 37.07 | <.001 |  |
| Insula | | | |
| T1w/T2w Ratio Quartile | 2.74 | .046 | 1^st^ and 4^th^ (*p*=.028) |
| Rasopathy Status | 37.54 | <.001 |  |

*via Tukey’s test

**Table S12. Effects of T1w/T2w ratios and RASopathy status in each region on PROMIS Strength ratings**

| Factor | *F* | *p* | Significant Quartile Differences* |
| --- | --- | --- | --- |
| Frontal Lobe | | | |
| T1w/T2w Ratio Quartile | 4.43 | .005 | 1^st^ & 3^rd^ (*p*=.034); 1^st^ and 4^th^ (*p*=.022) |
| Rasopathy Status | 9.38 | .002 |  |
| Parietal Lobe | | | |
| T1w/T2w Ratio Quartile | 6.33 | <.001 | 1^st^ and 4^th^ (*p*=.021); 1^st^ and 3^rd^ (*p*=.012) |
| Rasopathy Status | 6.57 | .011 |  |
| Temporal Lobe | | | |
| T1w/T2w Ratio Quartile | 4.20 | .007 | 1^st^ and 4^th^ (*p*=.013); 2^nd^ and 4^th^ (*p*=.037) |
| Rasopathy Status | 8.31 | .004 |  |
| Occipital Lobe | | | |
| T1w/T2w Ratio Quartile | 7.45 | <.001 | 1^st^ & 3^rd^ (*p*=.020); 1^st^ and 4^th^ (*p<*.001);  2^nd^ and 4^th^ (*p*=.006) |
| Rasopathy Status | 7.08 | .009 |  |
| Cingulate Cortex | | | |
| T1w/T2w Ratio Quartile | 5.14 | .002 | 1^st^ and 4^th^ (*p*=.002); 2^nd^ and 4^th^ (*p*=.041) |
| Rasopathy Status | 7.92 | .005 |  |
| Insula | | | |
| T1w/T2w Ratio Quartile | 3.54 | .017 | 1^st^ and 4^th^ (*p*=.037) |
| Rasopathy Status | 8.18 | .005 |  |

*via Tukey’s test

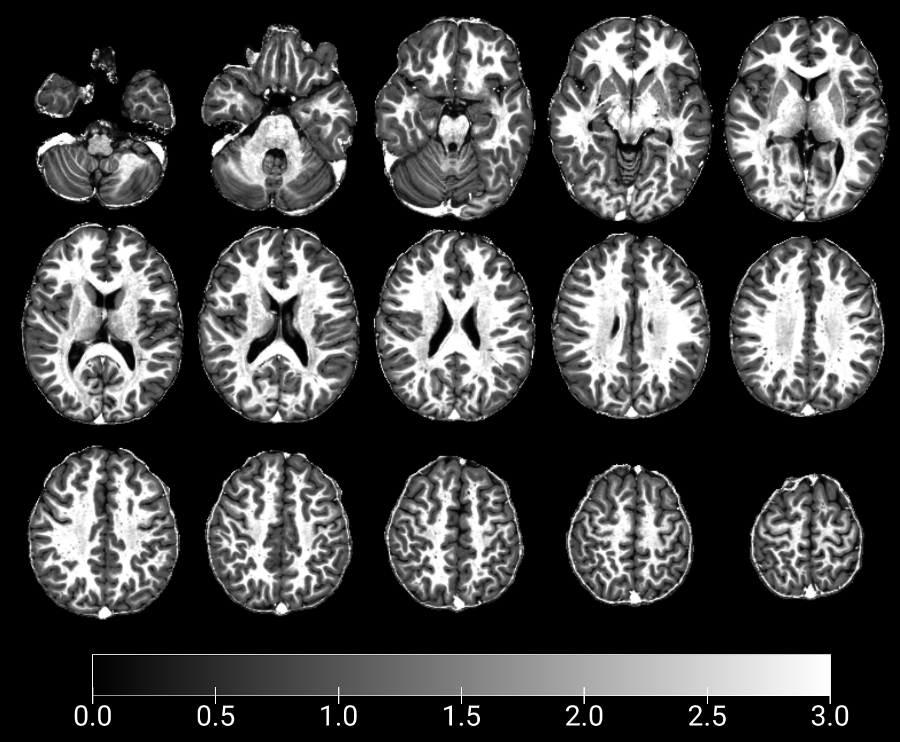

**Figure S1. T1w/T2w ratio maps of selected slices from an example participant.** Greyscale bar indicates T1w/T2w ratio signal from low (black) to high (white). Slices shown are (top row, from left) -40, -30, -20, -10, 0; (central row, from left) 10, 15, 20, 25, (bottom row, from left) 30; 35, 40, 45, 50, 55. Image created using MRIcroGL.

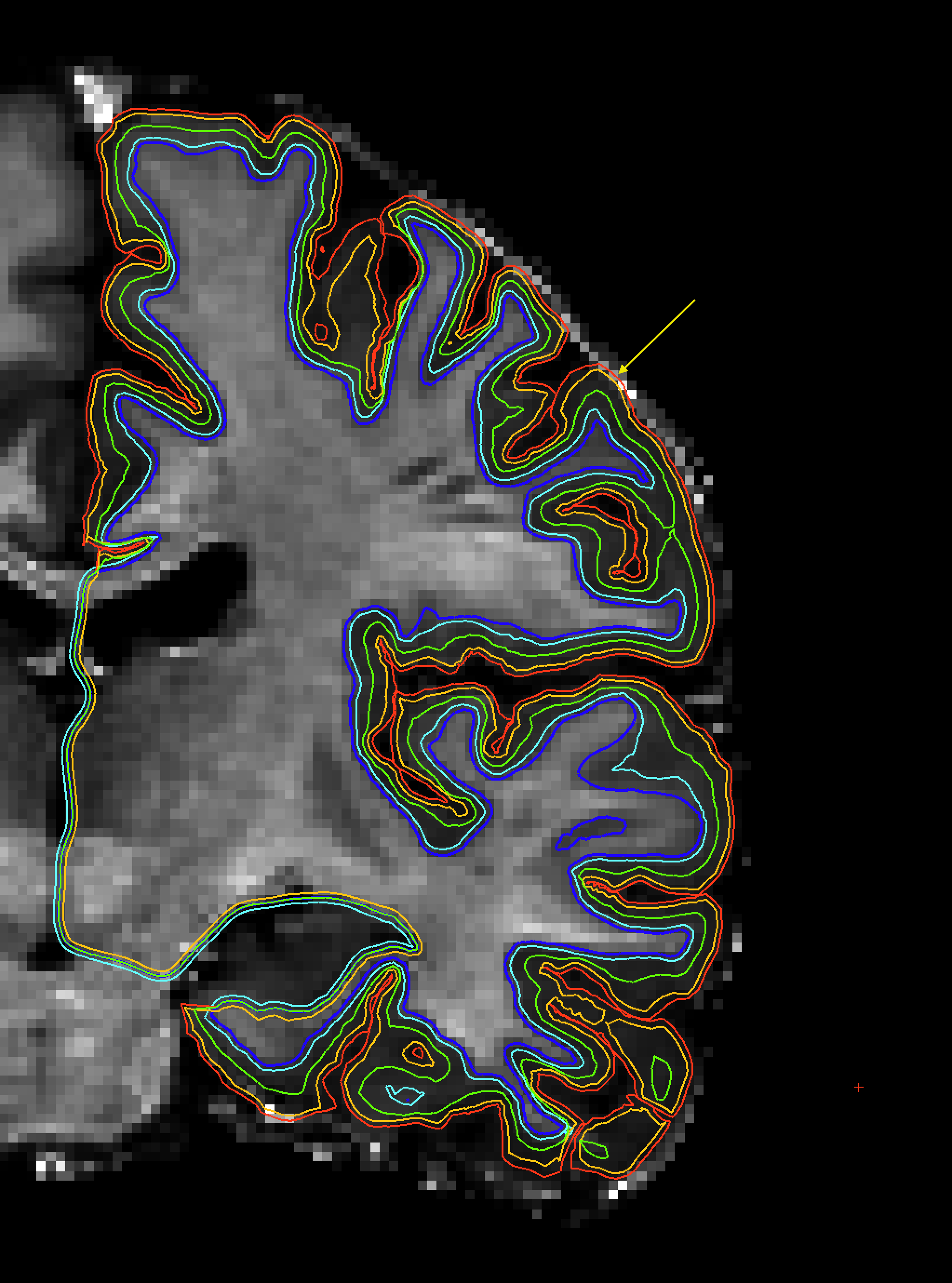

**Figure S2. Example coronal T1w/T2w ratio image showing a partial volume effect along the FreeSurfer-defined pial surface (red line). By projecting 0.5 mm below the pial surface (yellow line), the partial volume effect can be avoided.**

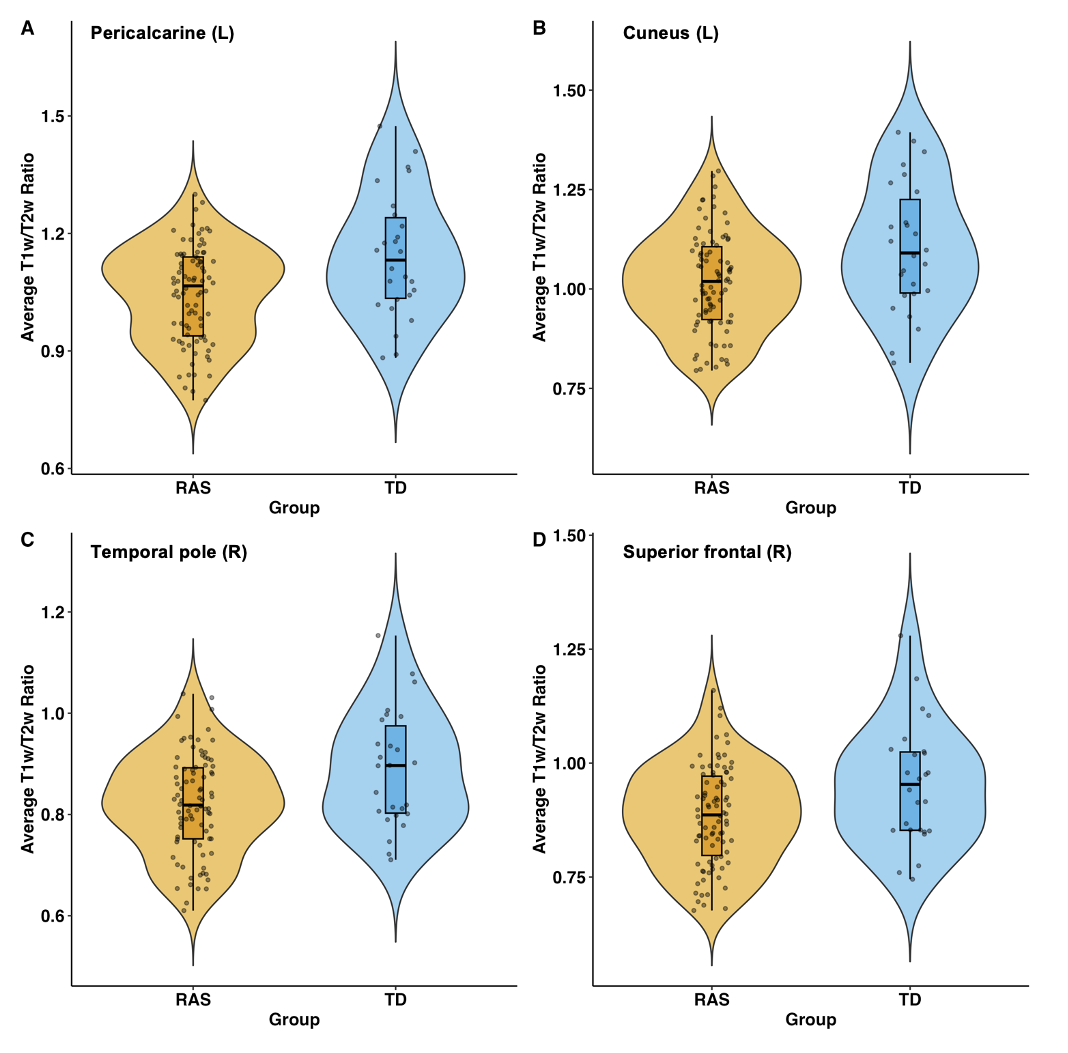

**Figure S3. Average T1w/T2w ratio comparisons between RASopathies and TD in selected cortical regions: A) left pericalcarine, B) left cuneus, C) right temporal pole, and D) right superior frontal.** All T1w/T2w ratio values shown were sampled from the midline. Each datapoint represents the average T1w/T2w ratio for an individual participant. All differences shown were significantly different between-groups (*p_FDR_*<.05). Figure created using *ggplot2* package in R v.4.4.1.

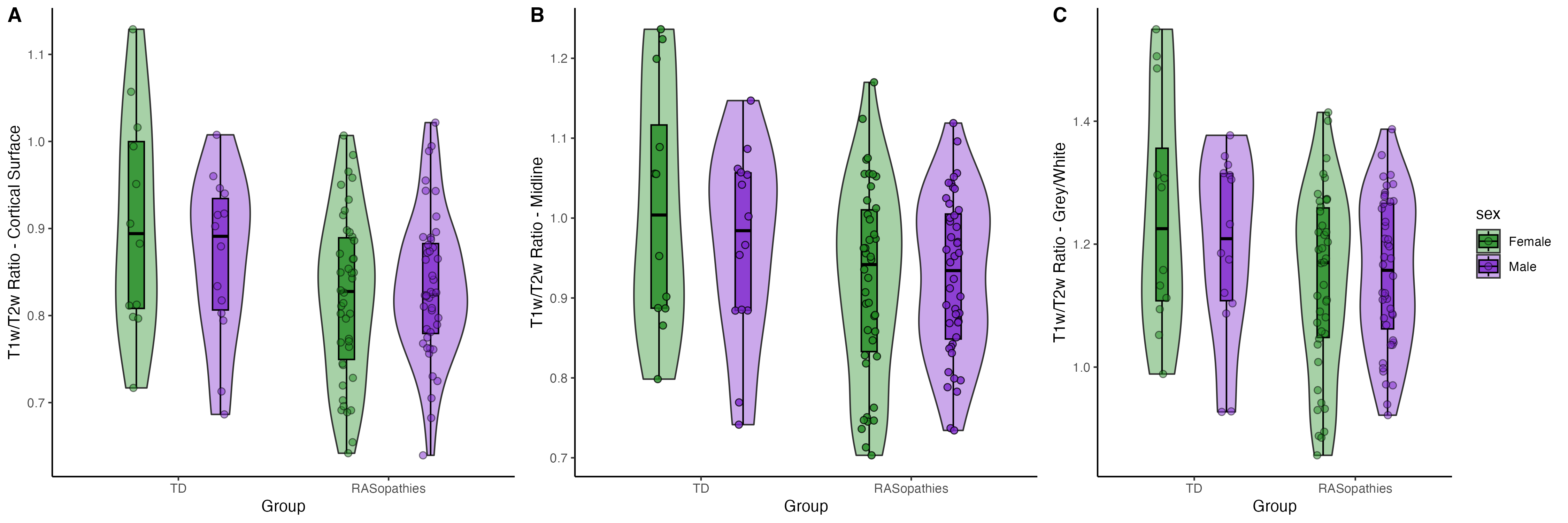

**Figure S4. Absence of sex differences in the average T1w/T2w ratios sampled from A) the cortical surface, B) midline, and C) grey/white boundary.** The T1w/T2w ratios were averaged across the 68 ROIs to generate an average ratio for each cortical level. Each datapoint represents the average T1w/T2w ratio for an individual participant. Figure created using *ggplot2* package in R v.4.4.1.

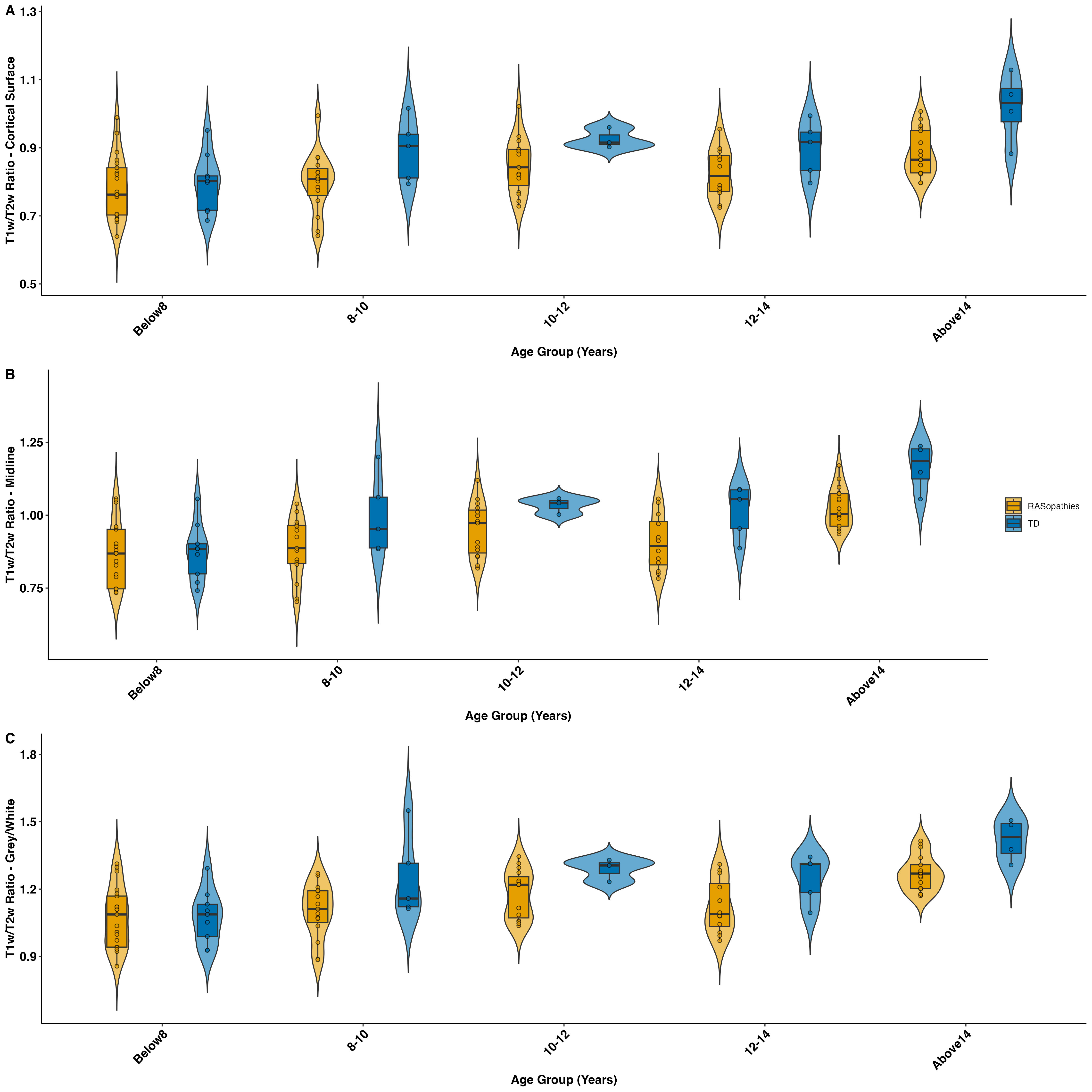

**Figure S5. The TD group shows a fairly consistent pattern of higher T1w/T2w ratios compared to RASopathies in each age group (below 8 years, 8-10 years, 10-12 years, 12-14 years, above 14 years) in each sampled cortical level A) cortical surface, B) midline, and C) grey/white boundary.** The T1w/T2w ratios were averaged across the 68 ROIs to generate an average ratio for each cortical layer. Each datapoint represents the average T1w/T2w ratio for an individual participant. Figure created using *ggplot2* package in R v.4.4.1.

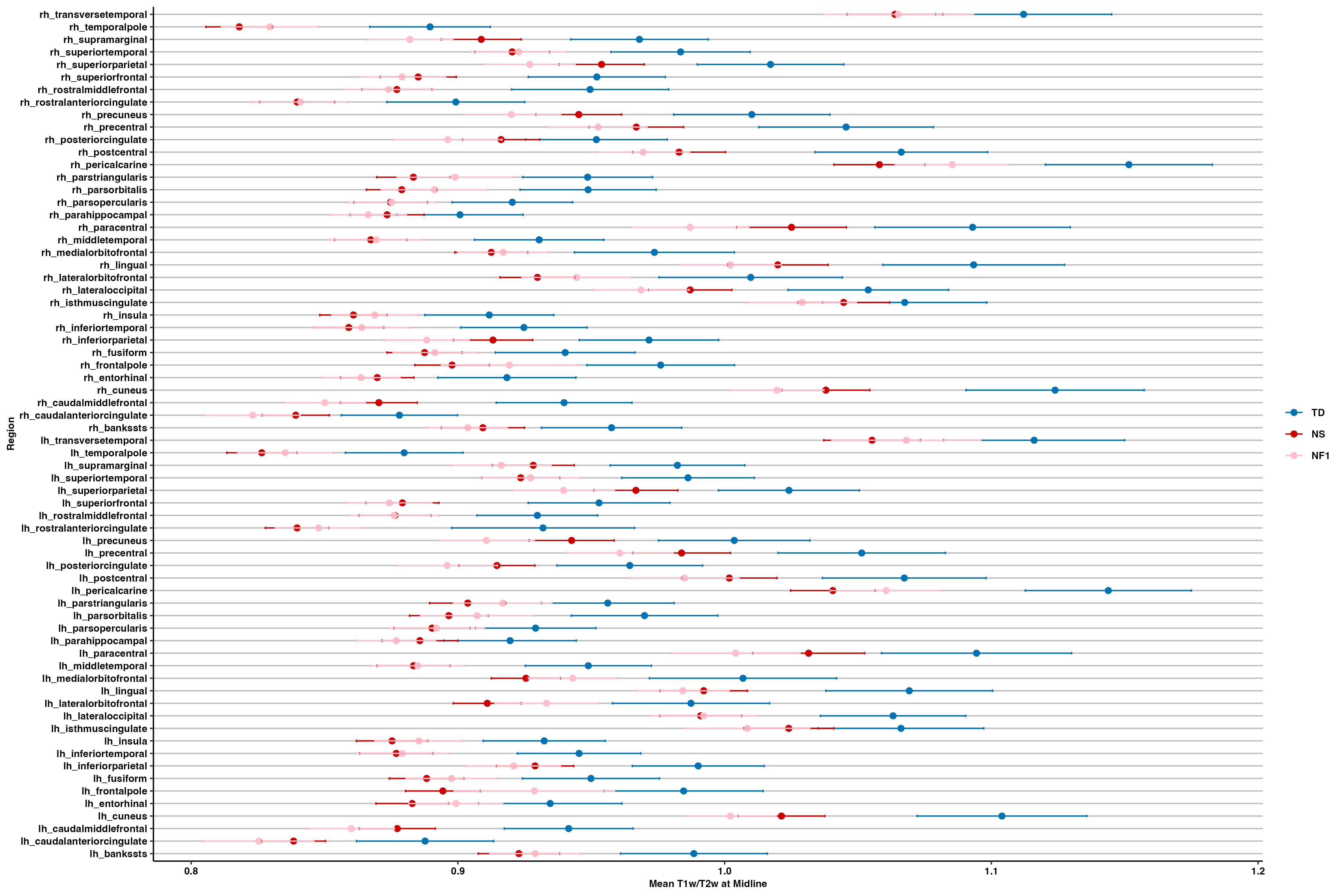

**Figure S6. Average T1w/T2w ratios and standard errors in cortical regions sampled along the midline in TD (blue), NS (red), NF1 (pink).** There were no significant differences between NS and NF1 in any of the cortical regions.

*RH=right hemisphere; LH=left hemisphere.*

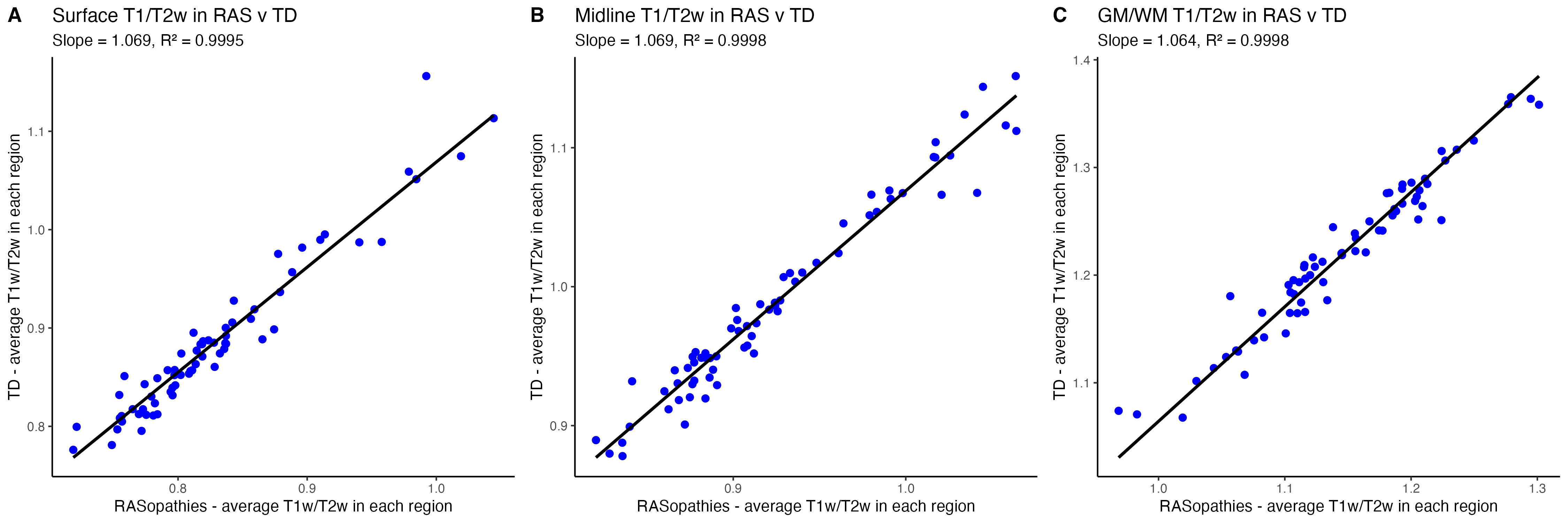

**Figure S7. Scatterplots showing the relationship between average regional T1w/T2w in TD (y-axis) and RASopathies (x-axis), based on T1w/T2w values sampled from the A) cortical surface, B) midline, and C) grey/white matter boundary.** Each datapoint (blue) indicates the average T1w/T2w value from one region. The black line is the line of best-fit estimated from each linear regression model.

*GM/WM=grey/white matter boundary; RAS=RASopathies; TD=typical developing.*

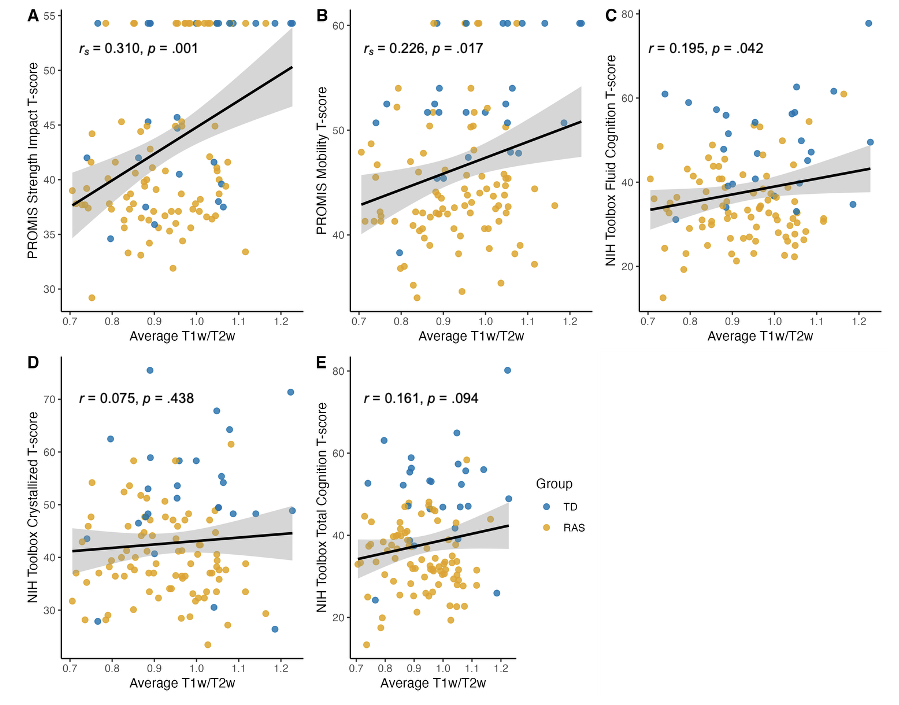

**Figure S8. Scatterplots showing the relationship between average whole-brain midline T1w/T2w and A) PROMIS strength impact t-score, B) PROMIS mobility t-score, C) NIH Toolbox fluid cognition t-score, D) NIH Toolbox crystallized cognition t-score, and E) NIH Toolbox total cognition t-score.** Correlations with PROMIS measures were conducted using Spearman, whereas cognitive correlations were conducted using Pearson. The black line is the line of best-fit estimated from each linear regression model and grey indicates the standard error.

*RAS=RASopathies; TD=typical developing.*

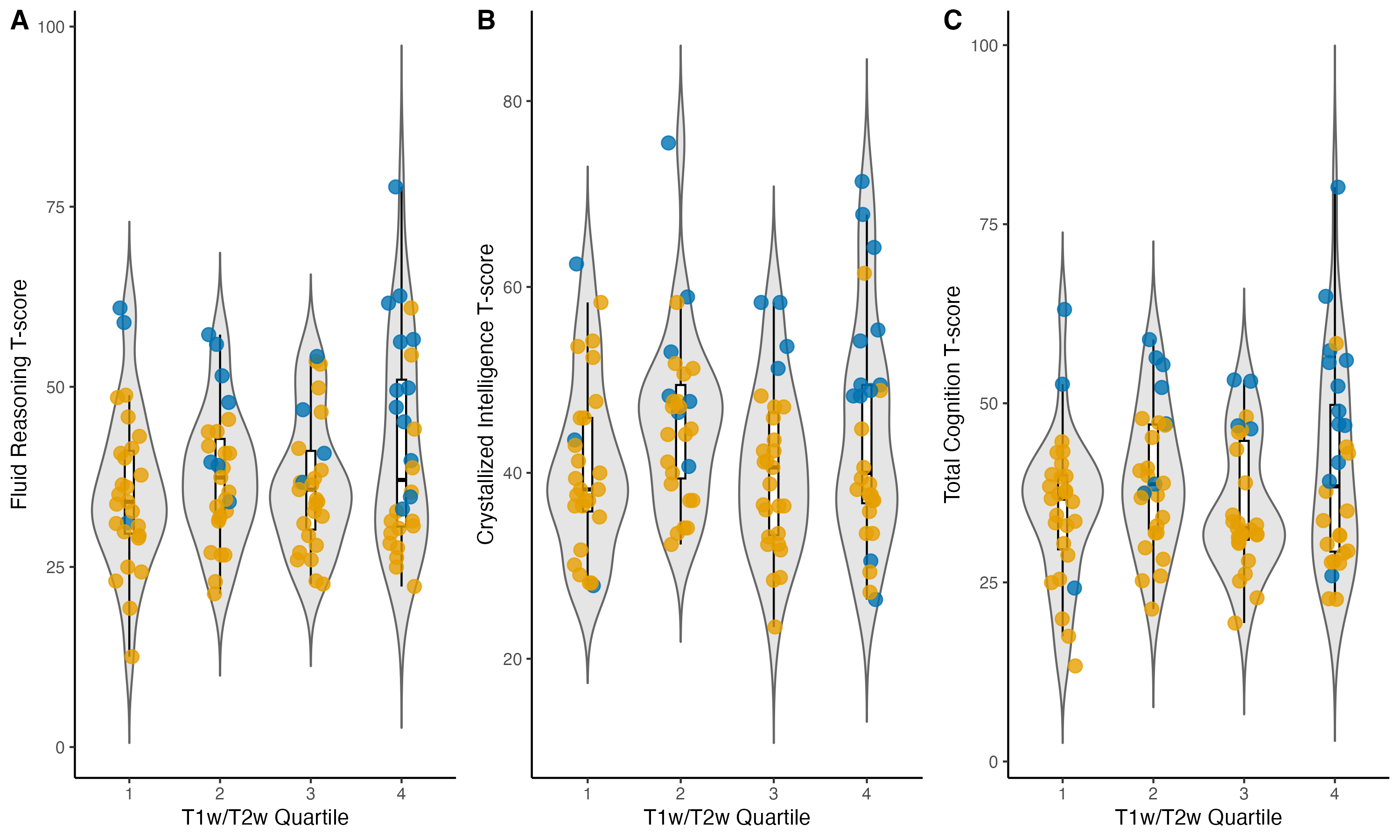

**Figure S9. Whole-brain T1w/T2w ratio quartiles according to A) fluid reasoning T-scores, B) crystallized intelligence T-scores, and C) total cognition T-scores, with datapoints color-coded by group.**

There were no significant differences in cognitive scores between the quartiles.

*RAS=RASopathies; TD=typical developing; T1w=T1-weighted; T2w=T2-weighted.*
